# A Healthcare Service Delivery and Epidemiological Model for Investigating Resource Allocation for Health: The *Thanzi La Onse* Model

**DOI:** 10.1101/2024.01.04.24300834

**Authors:** Timothy B. Hallett, Tara D. Mangal, Asif U. Tamuri, Nimalan Arinaminpathy, Valentina Cambiano, Martin Chalkley, Joseph H. Collins, Jonathan Cooper, Matthew S. Gillman, Mosè Giordano, Matthew M. Graham, William Graham, Eva Janoušková, Britta L. Jewell, Ines Li Lin, Robert Manning Smith, Gerald Manthalu, Emmanuel Mnjowe, Sakshi Mohan, Margherita Molaro, Wingston Ng’ambi, Dominic Nkhoma, Stefan Piatek, Paul Revill, Alison Rodger, Dimitra Salmanidou, Bingling She, Mikaela Smit, Pakwanja D. Twea, Tim Colbourn, Joseph Mfutso-Bengo, Andrew N. Phillips

## Abstract

**Background:** Decisions need to be made in all healthcare systems about the allocation of available resources with the aim of improving population health. Evidence is needed for these decisions, which can have enormous consequences for population health, especially in lower-income settings.

**Methods:** We address this need using an individual-based simulation model of healthcare need and service delivery that we have developed for Malawi, drawing on demographic, epidemiological and routine healthcare system data (on facilities, staff, and consumables). We compare the model’s simulated outputs with available data and estimate the impact that the healthcare system is having currently. We analyse the effects of improvements in healthcare access, clinician performance and consumables availability.

**Findings:** Malawi’s healthcare system averted 40 million Disability-Adjusted Life-Years (DALYs) in the five-year period to end-2019, which is half of the total DALYS that the population (total size: 19 million in 2020) would otherwise incur. This impact is strongly focussed on young children (mediated largely by programmes addressing respiratory infections, HIV/AIDS and malaria) and also by the HIV/AIDS and TB programmes (among adults). More services seem to be delivered than would be expected based on the number of staff and expected time needed for services. Nevertheless, the additional services that are provided (through service times being reduced or additional HCW hours worked) account for half the impact of the healthcare system (i.e., ∼20 million DALYS averted). If system improvements gave ill persons immediate access to healthcare, led to optimal referral and diagnosis accuracy, and eliminated consumable stock-outs, the overall impact of the healthcare system could increase by up to ∼30% (12 million more DALYS averted).

**Conclusions:** The healthcare system in Malawi generates substantial health gains to the population with very limited resources. Strengthening interventions could potentially increase these gains considerably and so should be a priority for investigation and investment. A detailed individual-based simulation model of healthcare service delivery is a valuable tool for healthcare system planning and for evaluating proposals for healthcare system strengthening.

## Introduction

In all countries, important decisions need to be made about how to allocate resources to collective healthcare provision. Health policymakers need to respond to questions such as which effective interventions to offer as part of its health service provision, and which must it forgo, to maximise health and reduce health inequalities? How should available resources be deployed and configured to achieve this? What would be the benefit of strengthening specific parts of the healthcare system and which should be prioritised? What future changes in the need for healthcare must be planned for, and what are the consequences of failing to do so?

These questions are fundamental to the operation of every healthcare system but they are especially important in low-income settings, where ineffective choices would represent much larger losses in health than elsewhere.^1,2^ This has been recognised at the highest levels of government in Africa: in February 2019, fifty-five Heads of State from across the African Union passed a resolution to reorient health spending and healthcare systems to target the diseases and conditions that have the greatest impact on mortality and human capital development, and sought technical expertise for doing so.^3^

There has been a rapid expansion in technical support: for instance, The Disease Control Priorities Network^4^ and the Tufts Cost-Effectiveness Registry^5^ provide evidence on intervention cost-effectiveness; the International Decision Support Initiative^6^ seeks to support the development of institutions; and the WHO have developed a healthcare system planning tool (‘*One Health Tool*’^7,8^). Although these approaches have marked a huge step forward, they do not fully respond to the pressing technical needs: databases of cost-effectiveness are challenged by results being incomparable and partial^4,9^; in cost-effectiveness analysis, the relevant value of the ‘cost-effectiveness threshold’ is not known^10^; and although the *One Health Tool* represents several diseases and interventions, they are considered independently and estimates do not represent healthcare production or fragilities in healthcare systems. None of these approaches is adequate by itself to guide the long-term redesign of the healthcare system, nor is adapted to the wealth of epidemiological and systems data that are increasingly available.^11^ Consequently, an extremely high value is attached to developing a new approach.^12,13^

We present an innovative approach in the form of a detailed individual-based simulation model, rooted in country-specific data, that represents each step in the generation of health gains delivered by the healthcare system. The model captures (i) the availability of resources to deliver healthcare services; (ii) the need for healthcare, as it arises from the multiple causes of ill-health that affect each person individually over their life; and (iii) the effectiveness of the care that is provided, accounting for the range of services that can be offered, the ability of persons to seek care, and the quality of care provided. We use the model to characterise how resources are used currently in Malawi to generate health gains in the population, and to quantify the impact of overcoming some of the frailties in the system.

## Methods

The scope of the model is illustrated in Figure 1. It aims to represent the capacity of the healthcare system to deliver healthcare services, the need for such services as they arise from causes of ill health, and the health consequences of receiving care (or not). These are represented as three interlocking sets of modules, briefly described in turn below. Full technical write-ups of all parts of the model^15^ and the source code are publicly available^14^; in addition, key components of the model have been described in earlier publications (cited below).

**Figure 1:**
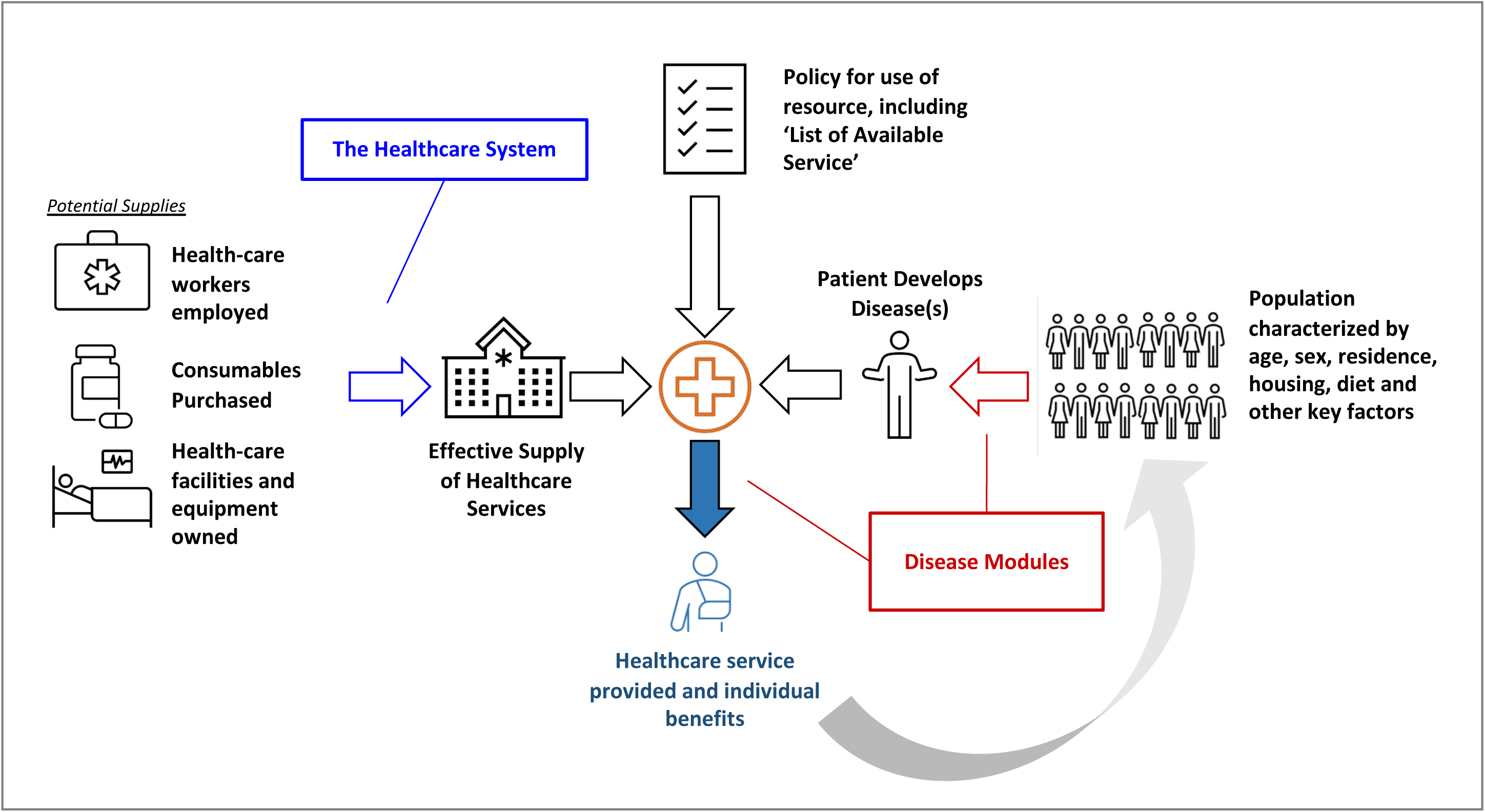
Schematic of model design. The overall model comprises: (i) a representation of how resources (health-care workers, consumables and facilities and equipment) combine to generate an effective supply of healthcare services (left-hand side); (ii) a representation of the population served by the healthcare system and the needs for healthcare that arise (right-hand side); and (iii) a set of policies that govern how resources for healthcare are marshalled to meet these healthcare needs (top). The grey arrow indicates the feedback that is incorporated between healthcare that is received (or not received) on the characteristics of the population and patterns of healthcare need in the future.

### 1) The Capability of the Healthcare System to Deliver Services

As described by She *et al.*^15^, there are four levels of healthcare facilities in the model (0 – community; 1 – primary care; 2 – regional referral hospitals; 3 – national referral hospitals). Facilities are specific to each district or region, and it is assumed that resources are pooled among all facilities at level 0 in the same district, among facilities at level 1 in the same district, among level 2 facilities in the same region, and among all level 3 facilities nationally. Each set of facilities is endowed with a complement of Healthcare Workers (HCW), of which there are 9 cadres (Clinical, Community Health Worker (CHW), Dental, Laboratory, Mental health, Nursing/Midwifery, Nutrition, Pharmacy, Radiography). Each cadre of HCW is assumed to be able to deliver healthcare services for a certain number of minutes per day, which is derived from the number of staff employed, their contracted hours and expectations for absences and leave^16^.

Healthcare services are delivered during what are termed *Healthcare System Interaction* (HSI) events that are characterised as occurring at a particular facility level and requiring a certain number of *appointments*. Appointments define a unit of care delivered by one or more HCW cadres. For example, one HSI may require 1 x ‘Antenatal Care Appointment’ at a primary care facility, which implies a requirement for 1, 11 and 0.4 minutes of time from a clinician, nurse/midwife, and pharmacist, respectively. Following Berman *et al.*^16^, fifty different types of appointment are defined, which can be grouped into broad categories: Consultation with DCSA [a community healthcare worker], Dental, General Inpatient/Outpatient Care, HIV Clinic, Laboratory Services, Mental Health Clinic, Nutrition Clinic, Pharmaceutical Dispensing, Radiography, RMNCH (Reproductive, Maternal, New-born and Child Health), Tuberculosis Clinic, and ‘Other’.

During an HSI event, consumables (e.g., medicines) may be required and the model tracks whether those consumables are available at each facility level (in each month and in each district) based on empirical estimates of stock-out frequencies^17^.

### 2) Need for Healthcare Services

#### Contraception, births, and lifestyle

Each person in the model has certain properties (e.g., sex, district of residence, wealth quintile, smoking status, body mass index), the distribution and evolution over time of which are determined primarily with reference to the Demographic and Health Survey (DHS) data.^18^ The method of contraception (or none) used by each woman is also simulated so as to match the detailed contraception calendar data in the same source.^19^ Together with estimates of fecundity and the effectiveness of the contraceptives, this determines the chance of conception for each woman. The process of pregnancy, labour and delivery are modelled mechanistically and calibrated to match available data^20^ (Figure S1).

#### Healthcare Seeking Behaviour

When a person develops the symptoms of ill health or becomes pregnant, they may seek healthcare, either immediately or after a delay, at a particular facility level in their district and region. In the model, the probability of care seeking depends on the symptoms as well as the person’s sex, age, wealth and residence (whether urban or rural), based on empirical estimates by Ng’ambi *et al*..^21–23^ If a person never seeks healthcare, then no healthcare is provided.

#### Mechanistically Modelled Causes of Death and Disability

Mechanistic models were developed for the causes of death and disability that collectively account for ∼81% of the deaths and ∼72% of the disability-adjusted life-years (DALYs) estimated to have been incurred in Malawi in the period 2015-2019 according to the Global Burden of Disease estimates (GBD)^24,25^ (Figure S2). The disease models can interact with one another and the underlying properties of each person, to represent, for example, the risk of several diseases being determined by the same risk factor, and one disease increasing the risk of acquisition or severity of another. The model representation of each cause has been carefully calibrated to data specific to that cause, often with additional guidance from program managers and disease experts.^18^

The list of causes modelled mechanistically is as follows (and full details of each are given elsewhere^18^): Neonatal Conditions (Neonatal Sepsis, Complications of Prematurity, Neonatal Encephalopathy), Conditions of Early Childhood (Acute Lower Respiratory Infection (ALRI), Diarrhoea, Undernutrition leading to stunting), Maternal Conditions (Obstetric Haemorrhage, The Hypertensive Disorders of Pregnancy, Maternal Sepsis, Obstructed Labour and Uterine Rupture, Gestational Diabetes, Anaemia in Pregnancy), Communicable Diseases (HIV, Tuberculosis, Malaria, Measles, Schistosomiasis), Non-Communicable Diseases (Cancers (Bladder, Breast, Oesophageal, Prostate and ‘Other’), Cardiometabolic Disorders (Diabetes Type II, Hypertension, Stroke, Ischemic Heart Disease, Myocardial Infarction), Chronic Kidney Disease, Chronic Lower Back Pain, Chronic Obstructive Pulmonary Disease, Depression, Epilepsy) and Injuries (Road Traffic Injuries, fatal and non-fatal Self-Harm).

#### Non-Mechanistically Modelled Causes of Death and Disability

Other causes are modelled collectively in a phenomenological manner that leads to an additional risk of death that matches to the age, sex, and period-specific GBD estimates of deaths from these causes. In addition, generic symptoms that may arise from these causes can occur and, as with other symptoms, may lead to healthcare seeking.

### 3) Delivery of Healthcare Services

The full list of HSIs, which represent the care that is required for each of the causes of ill health and disability that are modelled mechanistically (according to Malawi Clinical Guidelines^26^), are given in Table S1. These are organised into streams of treatments (*TREATMENT_IDs*) that group HSIs that provide a particular treatment for a particular condition (Figure S3).

When healthcare is needed, the corresponding HSI for that individual is entered into a queue at the relevant facility level and district. Each day in the simulation, and for each queue, the total call on HCW time is compared to the time available. Where there is sufficient time for each healthcare worker to meet all the calls, then all the HSI events are run. Where there is insufficient time for at least one type of HCW, then two assumptions are possible: (i) “Elastic Constraints”, whereby all HSIs run (but implicitly, the time allocated to each is reduced and/or the time used for HCW is increased); or (ii) “Hard Constraints”, whereby some HSIs (those ascribed with a lower priority) are postponed to the next day or do not happen at all.

When an HSI event runs, consumables used during the treatment are logged and the beneficial impact of the treatment provided is simulated. If the effectiveness of that treatment is contingent upon a consumable item (e.g., a medicine) that is not available at the time at the facility, then the beneficial effect of the treatment is not simulated and the HSI event may or may not be rescheduled (depending on the healthcare seeking behaviour assumptions made for that HSI).

The various appointment types can be requested through multiple streams of treatment (Figure S4), all of which draw on the same available time of HCW, and all HSI events are subject to the same pattern of consumables availability. Consequently, as intended, the model effectively represents all the different types of care using the same finite resources.

### Computational Approach

The model is an individual-based simulation in the Python programming language^27^ and uses the pandas data analysis library.^28^ Each disease, demographic and behavioural process that is modelled is encapsulated with a code *Module*, which contribute ‘events’ to a queue (Figure S5). The simulation proceeds by executing these events in chronological order. Events can read from and write to a centralised store containing characteristics of individuals in the population. HSI are a specialised category of events that run subject to checks on the healthcare resources required.

For the analyses presented, the simulation starts on 1^st^ January 2010 with a representative model population size 100,000. All results are scaled to reflect that 1 person in the simulation represents approximately 145 real persons in Malawi. The central estimate and uncertainty range of any computed statistic is the mean and 95% interval, respectively, across ten runs of the model with the same parameter values. We have found this gives stable results that are comparable with those for larger population sizes and more runs.^19^

### Model Outputs

The model logs the date and nature of all demographic, disease-related events, and HSI events. The DALYs that are incurred in the population are computed as the sum of Years of Life Lost (when a death occurs, the number of life-years lost is equal to the difference between the age at death and a reference value for life-expectancy^29^) and Years Lived With Disability (the weights that attach to each health condition are those from Salomon *et al.*^30^; the disability for each individual from all conditions is measured monthly and combined additively).

### Analysis Plan

The central aim is to examine the relationship between the way in which available resources for health are used and the health impact achieved at a population level. First, in order to establish that model represent the “status quo” in respect of the epidemiology of the major causes of death and disability and the current usage of healthcare resources, outputs of the model were compared with available data in Malawi for the five-year period 2015-2019 (the most recent five-year period for which there is complete data from all relevant sources and prior to the exceptional events during the Covid-19 Pandemic (the impacts of which are described elsewhere^31^). Next, the model was used to estimate the current impact of the healthcare system, by making a comparison between the estimated population health under the status quo scenario and a counterfactual scenario of no healthcare being provided. Finally, the model was used to quantify the potential health gains that may be possible from specific parts of the system being strengthened (those in respect of improved patient healthcare seeking and access, improved clinical practice, and improved availability of consumables), by representing the effects of such changes as new scenarios (Table 1) and comparing the population health gain achieved to that under status quo.

**Table 1:**
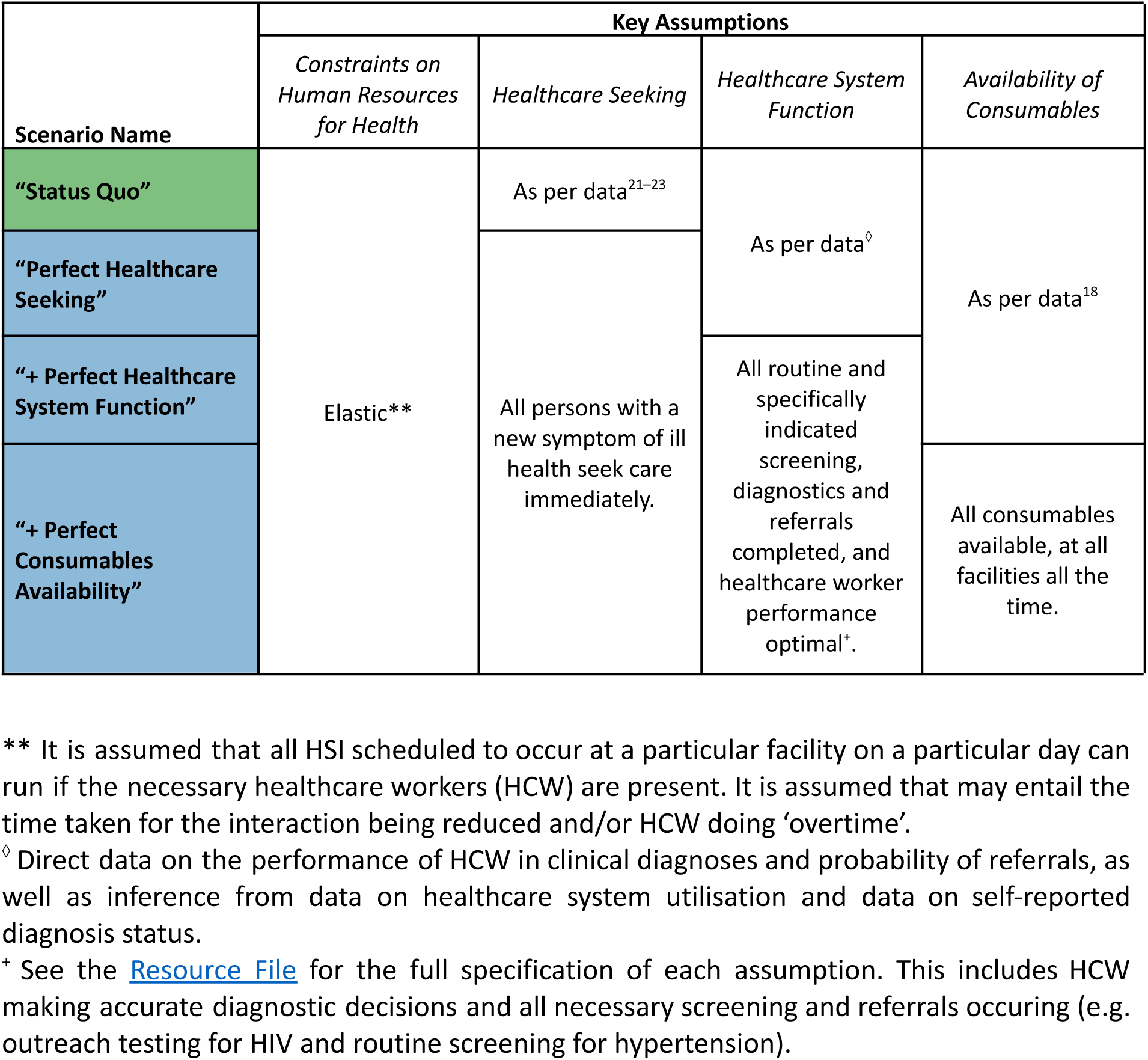
The Scenarios Used To Estimate the Health Impact of Healthcare System Strengthening.

## Ethics

The *Thanzi La Onse* project received ethical approval from the *College of Medicine Malawi Research Ethics Committee* (COMREC, P.10/19/2820) in Malawi. Only publicly available anonymised secondary data is used in the *Thanzi La Onse* model; therefore, individual informed consent was not required.

## Results

### (i) Representation of the ‘Status Quo’ in Malawi

#### Comparisons to Demographic and Epidemiological Data

The demographic outputs of the model were compared with available data (Figure S6). This shows that the population size growth, population pyramid, number of live births and the distribution of ages of mothers, all match closely with available data (the 2018 Malawi Census^32^ and the World Population Prospect (WPP)^33^).

The total number of deaths that occur in the model were compared with available data. The model provides a close match to available data (GBD^24,25^ and WPP) for the total number of deaths overall and when broken down by sex and five-year age-group (Figure 2(A-B)).

**Figure 2:**
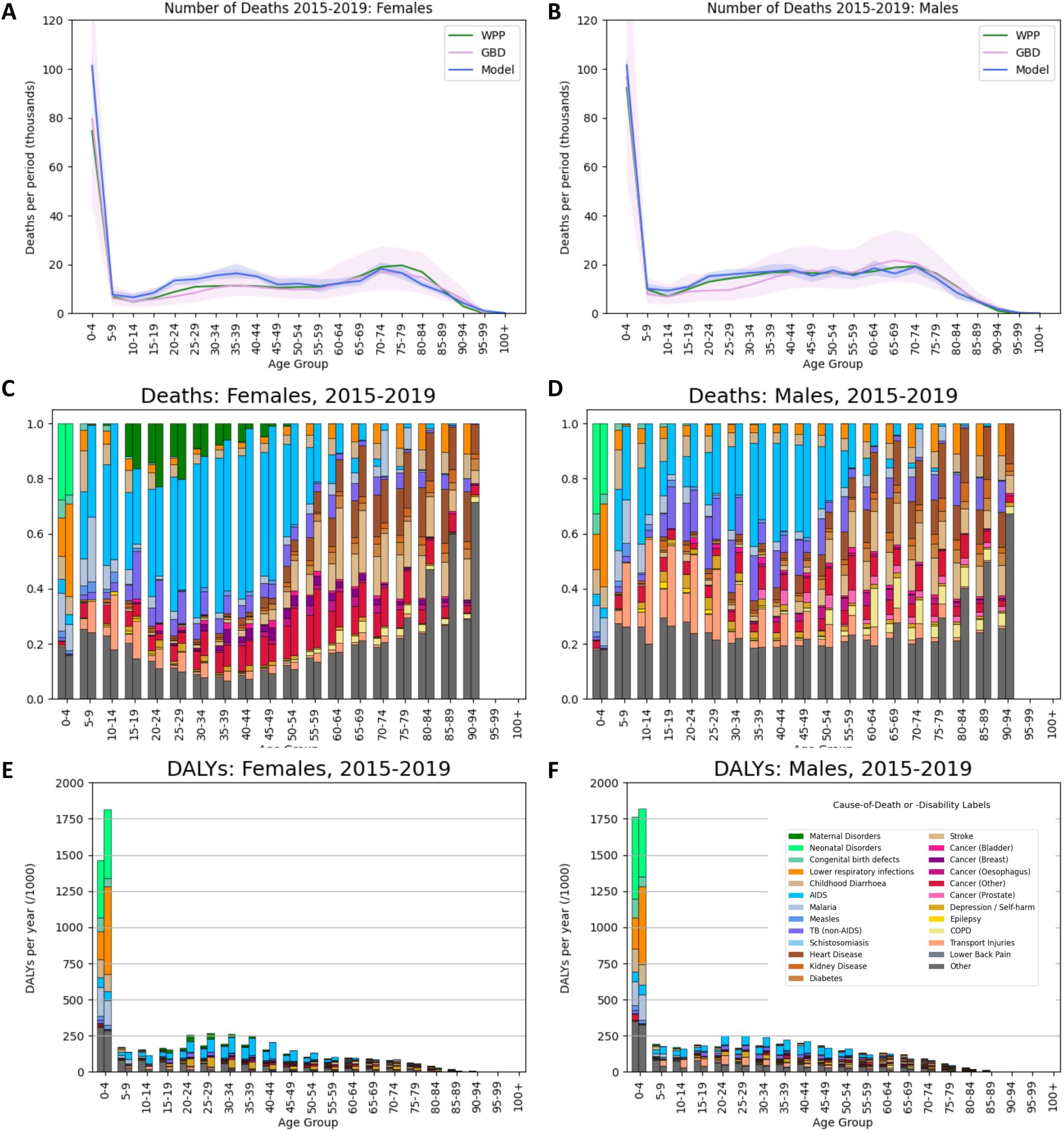
The burden of diseases in model projections in the period 2015-2019. (**A** & **B**) The total number of deaths by age-group in the model (blue line with shaded 95% uncertainty interval) and in the estimates by the Global Burden of Disease (GBD; pink line with shaded uncertainty interval) and World Population Prospect (WPP; green line with shaded uncertainty interval) for females (A) and males (B). (**C** & **D**) The causes of death by five-year age-group in the model (right-hand bars) and in the estimates by GBD (left-hand bars) for females (C) and males (D). Each bar shows the fraction of deaths in that age/sex group that is attributed to each cause, and ‘Other’ means causes that are not represented mechanistically in this model. (**E** & **F**) The DALYS incurred for each cause by five-year age-group in the model (right-hand bars) and in the estimates by GBD (left-hand bars) for females (E) and males (F). For panels C-F, the colour-coding of the causes is given in the legend inset in (F).

Each cause of death and disability in the model has been compared to data from disease-specific studies^18^, but it is also instructive to examine the distribution of deaths and DALYs that occur across the population. This distribution is not well characterised in Malawi but one source of estimates is the GBD. The model output for the distribution of causes of deaths and DALYs is in good agreement with the GBD estimates (Figure 2(C-F); Figure S7). The discrepancies that are apparent between the model and these data are mostly for Transport Injuries and, Depression/Self-harm and HIV/TB, and these are domains for which the model’s output match national data well.^34,35^

#### Comparisons to Healthcare Service Data

In the model simulation for the period 2015-2019, most HSIs are attributable to initial triaging (‘First Attendance’ interactions), contraceptive services, routine immunisation and care needed for malaria, following road traffic injuries, and HIV and TB (Figure 3(A)). Correspondingly, the appointment types needed most frequently are: ‘RMNCH’, ‘Outpatient’, ‘Consultations with DCSA’, and ‘HIV/AIDS-specific appointments’ (Figure 3B).

**Figure 3:**
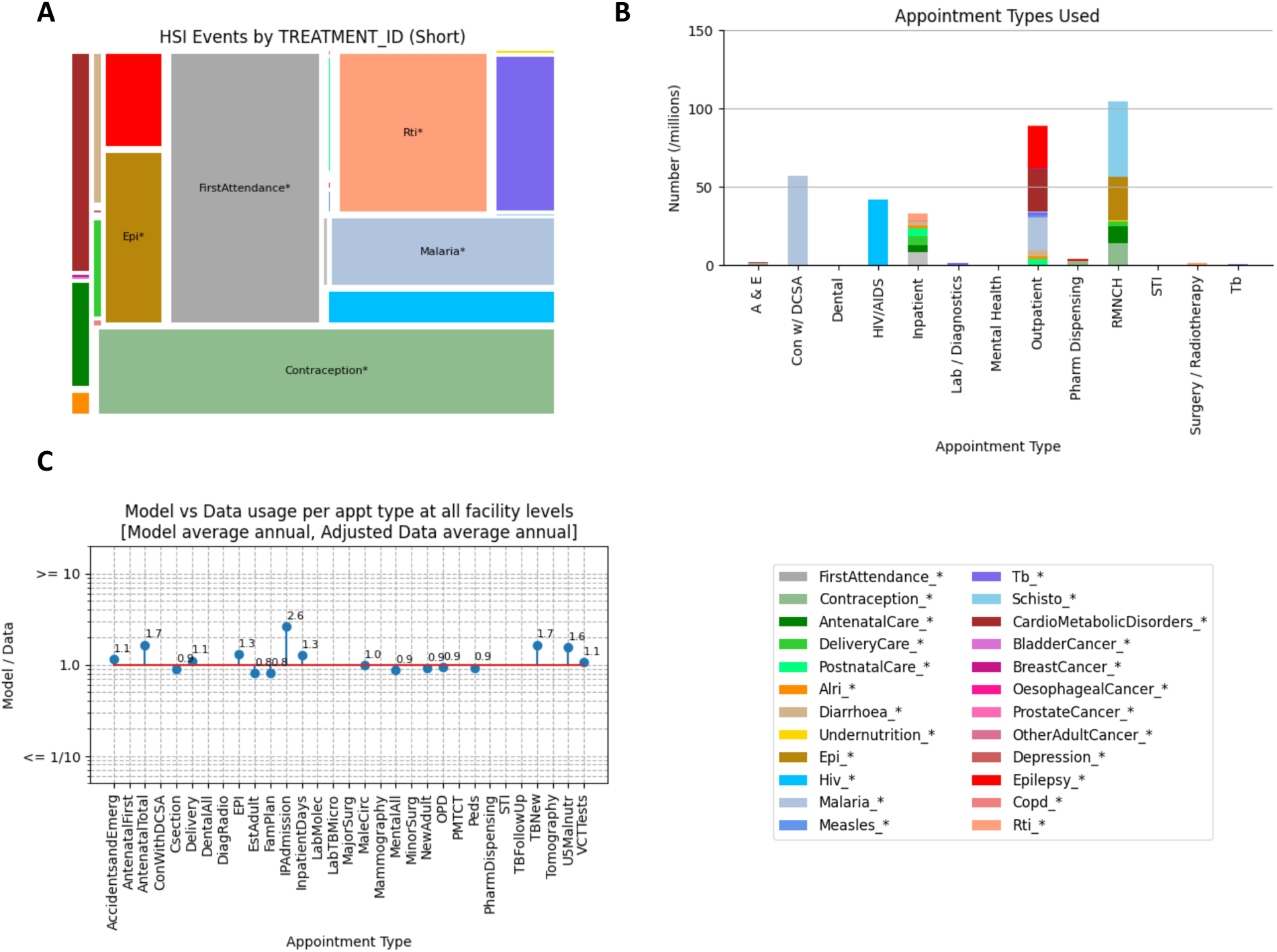
Modelled resource use in the Healthcare System in the period 2015-19. **(A)** The share of ‘Health System Interaction’ (HSI) events that occur in the model healthcare system broken down by the service provided (“TREATMENT_ID”). **(B)** The number of appointments of different types that occur for each service provided (TREATMENT_ID; indicated by the colour). (**C**) The “model: data” ratio of the total number of appointments per year of each type in the model to an analogous estimate from electronic records in Malawi.

This total number of each type of appointment delivered in this period were compared with records in the electronic medical records system^15,31^ (Figure 3(C)). There is a good match between the model and these data, with the ‘model-to-data’ ratio of frequency of appointments being close to 1.0 for most types, including the appointment types needed most. Some less common appointment types are outliers, with the ‘model-to-data’ ratio being above 1.0 (up to 2.6), which is consistent with the data slightly under-counting these types and/or the model overestimating their frequency. Although these data should in principle capture every type of appointment at every facility, some data are missing, and no data are captured at facility level 0.

### (ii) The Impact of Healthcare Services

The impact of all currently available services is estimated by comparing simulations of the “Status Quo” (as described above) and simulations in which no healthcare services are provided. In total, the healthcare system averted 40 million DALYs in the period 2015-2019, which is half of the 80 million DALYs that the population would otherwise have incurred.

The DALYs averted are skewed heavily to young ages, due to services averting DALYs that would be caused by ALRI, HIV/AIDS, malaria and Neonatal Disorders, in particular (Figure 4(A); Figure S8). The DALYs averted among adults would mostly have been attributed to HIV/AIDS and TB. The distribution of DALYs averted with respect to wealth quintile reveals a modest skew in favour of those in the highest wealth quintile compared to the average (Figure 4(B)), which is mostly attributed to wealthier persons having a great propensity to conditions that incur high morbidity/mortality, and, to a lesser extent, also being more likely to access healthcare ^21,23^ (Figure S9).

**Figure 4:**
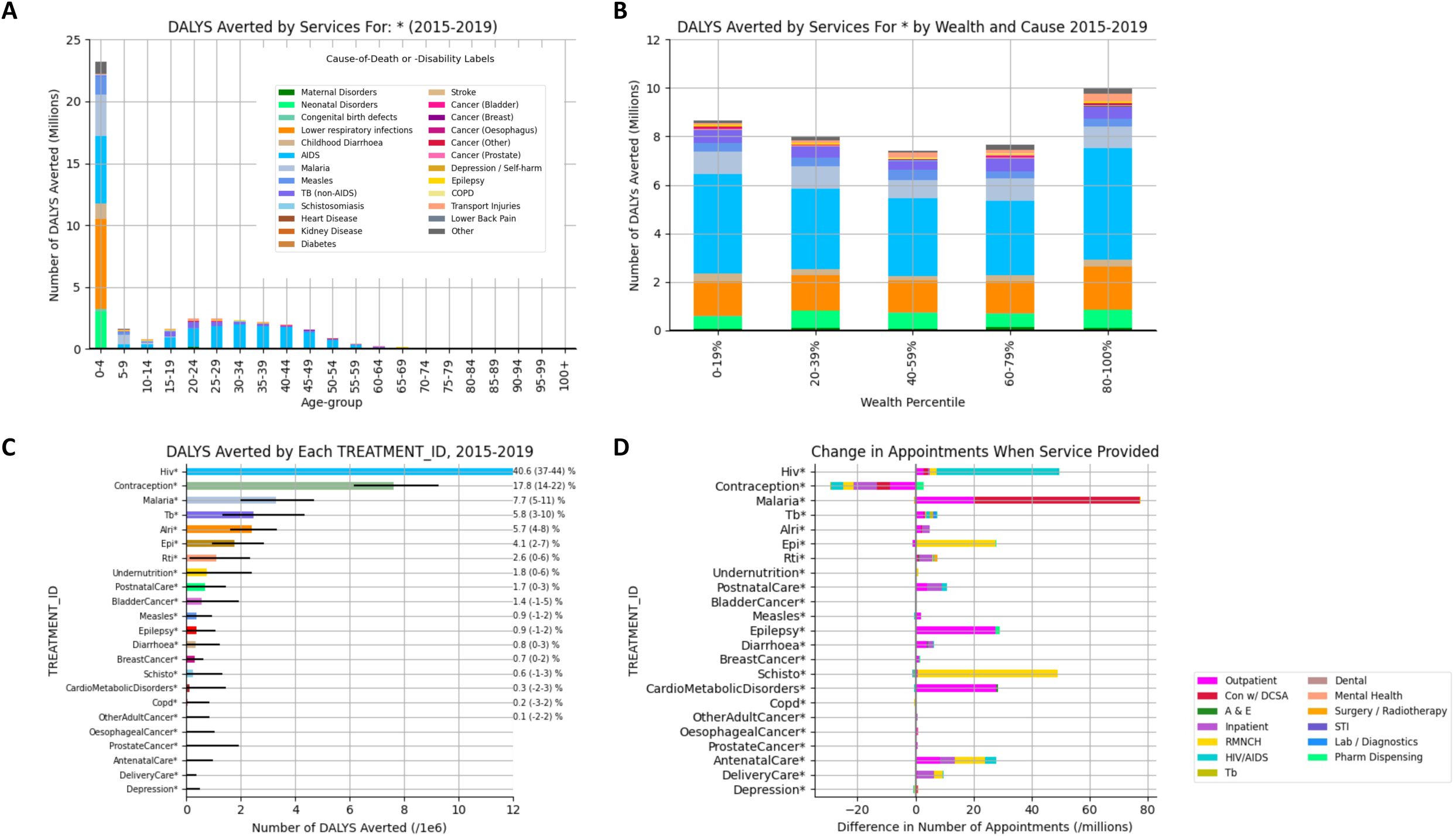
The modelled impact of healthcare services in the period 2015-2019. **(A & B)** Model projection of the disability-adjusted life-years (DALYs) averted by all healthcare services in Malawi, by five-year age-group (A) and wealth quintile (B). (**C**) The effect of each type of service (‘TREATMENT_ID’), expressed as the number of DALYs averted (bars) and as a percentage of all the DALYS incurred to when all interventions are available (annotations on right-hand side). (**D**) The difference in number of appointments needed when each services is provided. The bars in C & D are ordered from the top by the descending number of DALYS averted by that service. Error bars show the 95% uncertainty interval on model estimates.

The impact of a particular set of services can be estimated by comparing the Status Quo scenario to one in which all services are offered *except* those services (Figure 4(C)). This reveals that the service types with the largest incremental health impact (DALYs averted) include those for HIV/AIDS, contraception, malaria, TB, ALRI and routine vaccination. It is notable that the interaction between the diseases leads to the impact of a healthcare service being distributed across several causes of DALYs. For instance, HIV services lead to averted deaths due to childhood diarrhoea (for which HIV is a key risk factor), as well as to AIDS; and the health impact of contraceptive services is in averting deaths among 0–4-year-olds due to neonatal disorders, ALRI, malaria and childhood diarrhoea (Figure S10).

The difference in the number of appointments with and without a particular service being provided is also computed (Figure 4(D)). This shows that the various services are associated with substantially different magnitudes and types of calls on resources, which are not necessarily in proportion to the health impact generated. For instance, the appointments required for TB services are relatively few, even though the health gain they generate is among the greatest of all services offered; and, providing contraceptive services, which have a very large health impact, leads to many fewer appointments overall because there is a large reduction in those to do with care for neonates that offsets the need for Pharmacy Dispensing appointments wherein the women receive the contraceptives.

The Status Quo scenario also reveals the difficulties being faced by the system. There is a significant mismatch between the total amount of services provided and the time of HCW available and the time expected to be required to provide each service.^16^ This mismatch is quantified as a ‘Squeeze Factor’ (Figure S11), and is found to affect certain types of care to a much greater extent than others (for instance, emergency care and surgical services, for which relevant HCW are less available). In practice, this mismatch must be solved through suitably qualified staff working longer hours than expected, or appointments being briefer than expected, or both. When it is enforced in the model that each appointment takes the time expected and that HCW do not work more for longer than contracted (The ‘Hard Constraints’ assumption), the impact of the healthcare system is reduced substantially as fewer services can be provided: in that case, the system would avert only 20 million DALYs in 2015-2019 (Figure S12), which suggest that half of the current impact is derived through overwork and services being ‘squeezed’.

We also note that many HSI are frustrated by a lack of availability of consumables and that many of the most frequently needed items are not available on up to ∼40% of occasions (Figure S13). For acute conditions, this may mean patients do not benefit from the treatment, while for longer-term conditions it may lead to delays and/or additional HSIs.

### (iii) The Impact of the Strengthening the Healthcare System

The effect of different types of healthcare system improvement can be estimated (Table 1; Figure 5). Compared to the Status Quo scenario, if persons becoming ill would immediately access care, then 8% (3.6 [2.0-5.3] million) more DALYS could be averted. If, in addition, all referrals were successfully completed and diagnostic accuracy of HCW was as good as possible, then 20% (8.8 [7.0-10.2] million) more DALYS could be averted. And, if, in addition, consumables were always available, then 28% (12.3 [10.9-13.8] million) more DALYS could be averted. This also implies that, despite all the frailties in the system, its functioning is still 78% as impactful as an idealised system that provided the same set of services.

**Figure 5:**
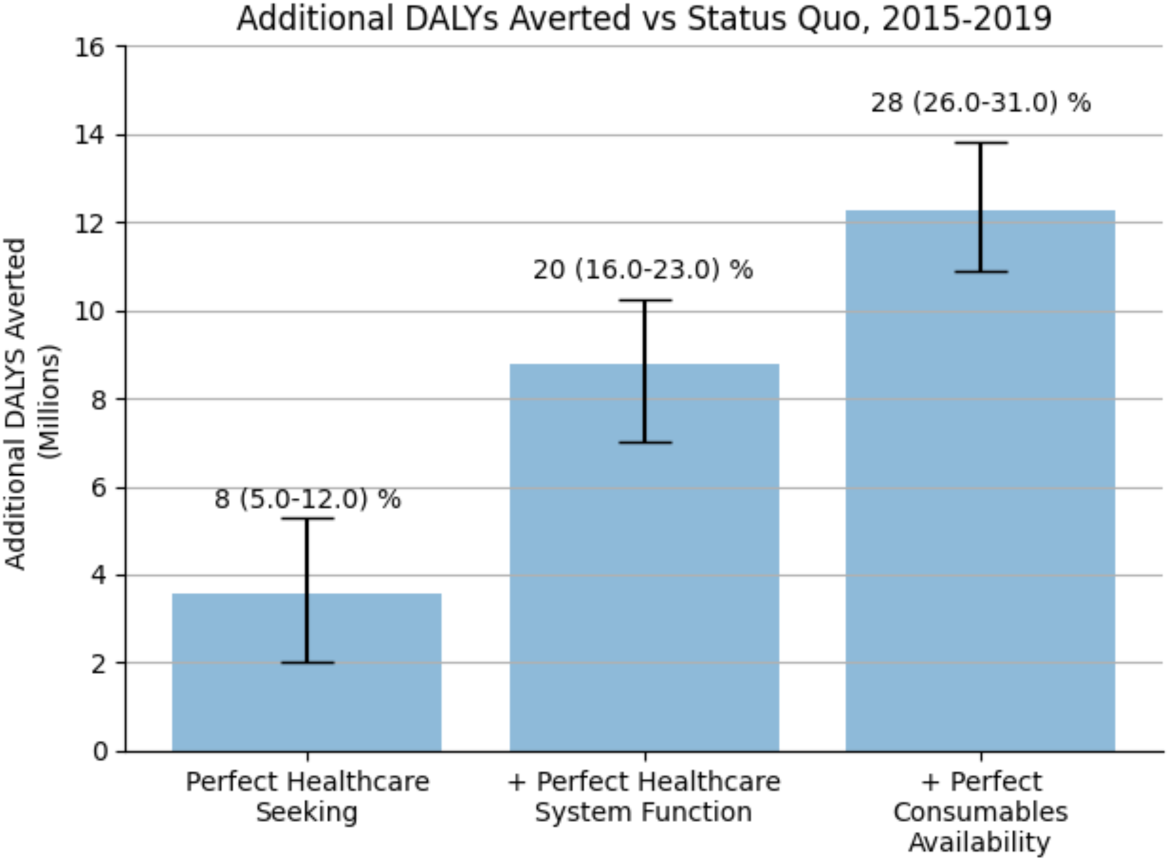
The Impact of the Strengthening the Healthcare System. The additional DALYs averted under each of the healthcare system improvement scenarios relative to the DALYs averted by the Healthcare System in the Status Quo scenario (per Table 1). Error bars show the 95% uncertainty interval on model estimates.

## Discussion

We have presented a new approach to answering fundamental questions about healthcare system service delivery and shown how it has been successfully applied to data from Malawi. In Malawi, this approach shows how the impact of the healthcare system is very substantial overall (50% DALYs averted in 2015-2019), strongly focussed on young children and, overall, is mediated largely by programmes addressing HIV, TB, malaria and ALRI, as well as contraceptive services. This analysis also reveals how the system operates with appointment times or HCW working hours far beyond what clinical expectations or contracts allow; but that without such flexibility (for reducing the time for appointments or overtime) the impact of the healthcare system would be only half as large. The consequences of other major frailties in the system have also been quantified – delayed healthcare seeking, imperfect referral completion and diagnostic accuracy, and consumable stock-outs – that, if overcome, could increase the overall impact of the healthcare system by almost 30%.

We believe that this data-grounded modelling approach will be foundational to addressing many issues that, hitherto, have not been amenable to such direct analysis. These applications include examining which services should constitute a ‘benefits package’ in view of the differential health gains and resource requirement of various services; quantifying the health gains (and so also the ‘Return-on-Investment’) of addressing weaknesses in many parts of the health system (e.g., through improved HCW availability, improved stock availability, redistribution of equipment to allow care to occur at different levels, and task-shifting such that different cadres can contribute to different services); and also revealing the “knock-on” consequences of interventions that are normally not quantified, such as how preventing disease not only benefits persons directly but also frees-up resources so that other forms of care can be provided. We therefore believe that our modelling approach lays the foundations for a new perspective on justifying and advocating for health system investment. Quantifying what can be achieved in terms of lives saved and disability avoided, across a whole health system, makes the tangible benefits of interventions clear and provides decision-makers with the evidence they need to persuade the financing agencies of the value of their contributions.

Our approach has several important strengths. By design it captures a large span of health conditions, transmission of infectious disease and development of conditions over the life-course, meaning that results automatically capture the indirect effects of different decisions (through interactions between diseases and underlying risk factors, and feedback via transmission), giving a fuller characterisation than would be possible under ‘siloed’ modelling approaches. The systematic integration of the available data for interventions, means that results that relate to different diseases or interventions are comparable, in contrast to results culled from literature that vary in terms of perspective, counterfactual and outcome measurement.^36^ Although the approach demands lots of data, these demands have been met in Malawi, and, we believe, will be met in other countries too. Furthermore, using data to inform decision-making in this way provides a key incentive for improving data systems.

The limitations to our approach include: (i) Not all causes of disease, nor all associated interventions, are represented mechanistically in the model, so the consequences of decisions for some diseases and interventions will not be quantified (the conditions to which this applies are shown in Figure S2). In addition, some of the diseases that are modelled explicitly have a very simplified description, which we consider to be sufficient for the purposes of these analyses, but which will not capture all possible interactions and calls on resources. We expect that disease models will be added, and others augmented, in this modular open-source software. (ii) It is not possible to fully validate the model because the data that would be required to do so do not exist (e.g., completely independent, reliable data on causes of death in Malawi; or, measured health outcomes from two comparator real-world healthcare systems that differ only in the frequency of consumables stock-outs). Furthermore, whilst we can infer that HSI are being ‘squeezed’, we cannot estimate the magnitude of relieving such an effect, even though one would presume it would improve the quality of care to some extent. Nevertheless, in Malawi, and elsewhere, some of these data gaps will be overcome, at least partly, through ecological comparisons between clinics with different management structures,^37^ or where the patient mix and volumes vary widely (studies are on-going). (iii) The fine detail of the model requires making some assumptions without the benefit of direct data: for instance, the behaviour of clinicians and patients when medicines are not available, or how much quicker healthcare seeking could become. Nevertheless, in other analyses these assumptions would still have been made implicitly and exposing them in this detailed simulation affords the possibility of investigating their effect and motivating further data collection.

We have found that, if contraceptive services were not available, ∼7.5million additional DALYs would have been incurred in the population (Figure 4(C)). This is a measure of ill-health experienced (by new-borns and their mothers) and proxies the care that would need to be met by the healthcare system. However, it does not weigh the utility that would also be associated with the additional births, nor the wider economic and demographic consequences.^19,38,39^ Given these deficiencies in the conceptualisation of DALYs, arguably it may be appropriate to compute the entire impact of the healthcare system as 33 million DALYS in 2015-2019 (rather than 40 million DALYS), which avoids counting the DALYS averted due to contraceptive services.

An holistic, integrated and dynamic approach to modelling service delivery by the healthcare system has long since been called for as a means of addressing analytically, and with the benefit of vast amounts of data, the major questions about options for healthcare system strengthening and resource allocation.^12,13^ We believe that this approach meets this need, does indeed reveal important new insights, and will open the way for myriad new analyses that will support decision making in the years to come.

## Data Availability

All data produced are available online at https://zenodo.org/records/10144016

https://tlomodel.org

## Acknowledgements

We thank our colleagues in the Government of Malawi Ministry of Health, including the Planning and Policy Development Department and acknowledge the expertise shared by Dr Rose Nyirenda, Dr Mike Chisema, Dr Lazarus Juziwelo, Dr Godfrey Kadewere, Prof Andreas Jahn, Benson Chilima and Damson Kathyola. We would also like to express our gratitude to the collaborators, attendees and organisers of the Kamuzu University of Health Sciences HEPU Think Tanks which have improved model design, development and analysis immeasurably. We thank Professor Edward Gregg (Royal College of Surgeons in Ireland, and Imperial College London) and Professor Amelia Crampin (Malawi Epidemiology and Intervention Research Unit, Kamuzu University of Health Sciences, and University of Glasgow) for useful discussions, especially about the cardiometabolic diseases part of the model. We thank Kate Bilsborrow for support in running the project.

## Contributions

The project was conceived and led by TBH and AP with critical inputs by JM, MC, TC, DM, GM, PT and PR. AT led the design of the modelling software. TBH, TM, AT, JHC, MG, EJ, BLJ, ILL, RS, EM, SM, MM, WN, BS, TC and AP primarily constructed the model, with key technical supervision and input provided by MS, NA, JC, AR, VC. AT, JC, DS, WG, MSG, SP, MG and MMG developed the modelling framework and associated infrastructure. The first draft of the paper was written by TBH and all authors reviewed multiple further drafts.

### Research in context

#### Evidence before this study

We searched PubMed on 6th November 2023 using the following search strategy: “((healthcare system OR health system) AND (low-income) AND (model*) AND (agent-based OR individual-based)”. We found 32 articles, most of which focus on the specific impact of one type of intervention for one disease, which is similar to what an earlier systematic review found.^40^ One article^11^ is a systematic review of models for health system research, which found a preponderance of studies in high-income settings focussed on acute care, elderly care and long-term care services, and argues that, “future work should now turn to modelling health systems in low- and middle-income countries to aid our understanding of health system functioning”. Another systematic review of “Health System Modelling”,^12^ also found no studies fulfilling the needs they describe, and concludes that there is an “opportunity to advance modelling methods to further understand the dynamics between health system inputs and outputs”. The need for the ‘whole health system’ perspective has also been identified specifically by Verguet *et al*.^13^.

#### Added value of this study

This is the first analysis that meets the need for “Health System Modelling” in a low-income setting and it accomplishes this using a novel individual-based modelling approach that spans most of the diseases faced in a population and most of the services delivered by the healthcare system. The application of the model in Malawi finds that the healthcare system reduces ill health in the population by about half but the system is severely overburdened. Interventions that strengthen service delivery and allow earlier healthcare seeking behaviour by patients could increase the health impact by up to 30%.

#### Implications of all the available evidence

Planning for healthcare service delivery can now benefit from analytic evaluations of alternative choices for service delivery and system strengthening interventions, wherein the real constraints and frailties in the healthcare system are reflected. This has been called for repeatedly and will provide crucial additional evidence to health policy decision-making and help raise and marshal resources to increase the beneficial health impact of healthcare systems.

