## Supplementary Figures for "A Healthcare Service Delivery and Epidemiological Model for Investigating Resource Allocation for Health: The *Thanzi La Onse* Model"

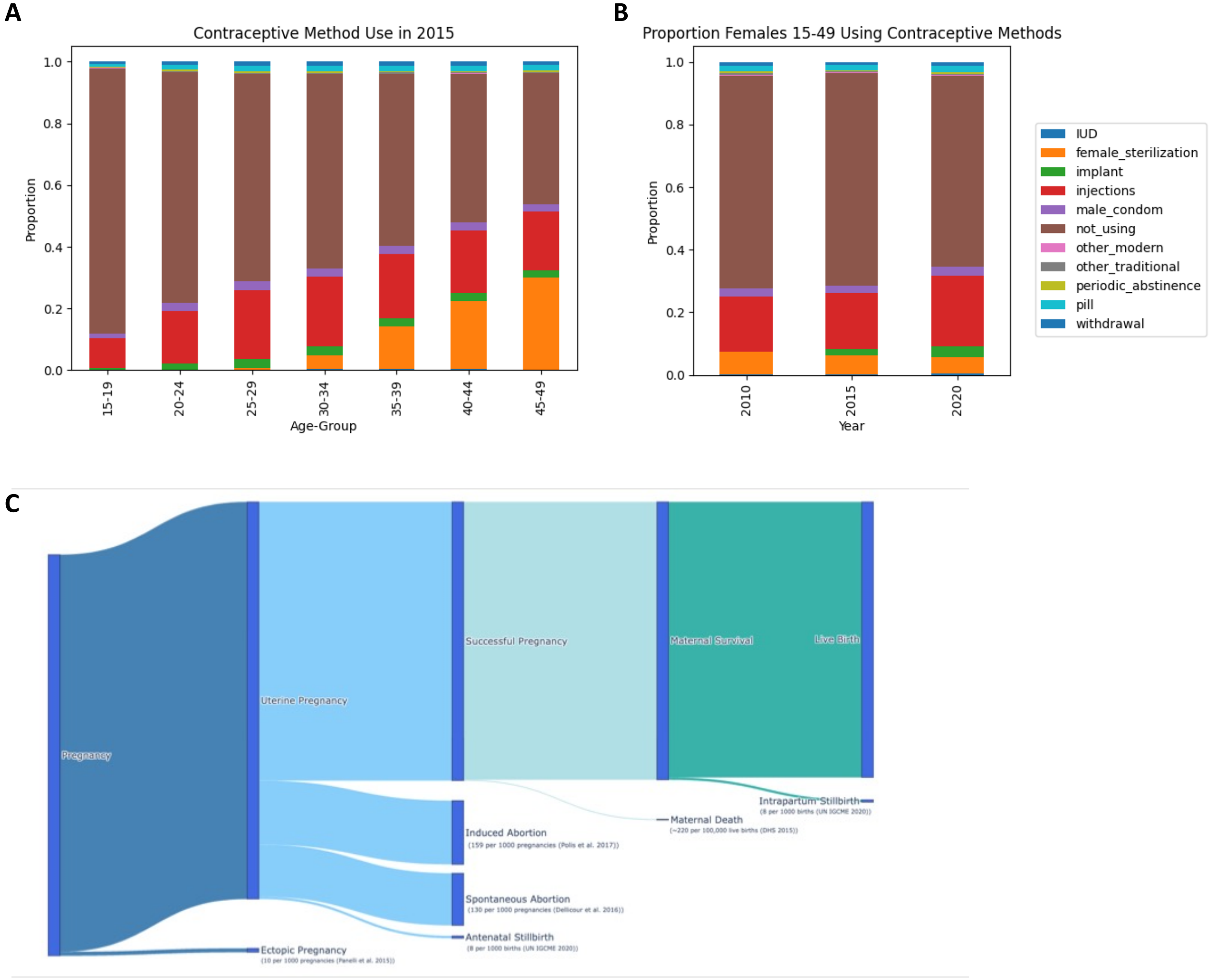


**<u>Figure S1</u>: The pathway to live births that is modelled.** (**A**) The distribution of use of contraceptives by women by five-year age-group in the model in the year 2015. (**B**) The proportion of women aged 15-49 using each type of contraceptive at the start of the year in 2010, 2015 and 2025. (**C**) The distribution of outcomes modelled for a hypothetical cohort of 1000 women following conception and through to live-birth. Where appropriate the estimated risk of each outcome is given with a suitable reference, which in each case is matched in the model.

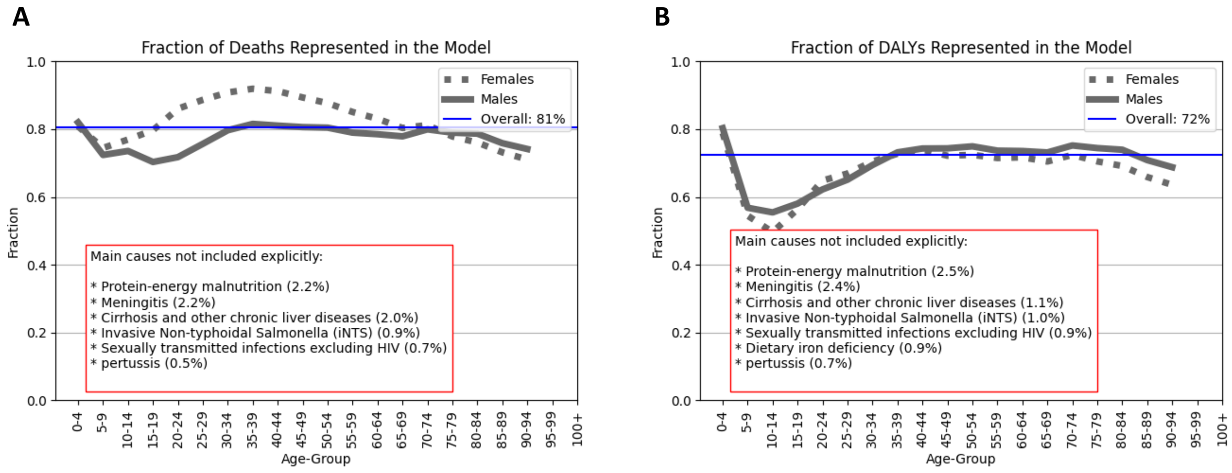


**Figure S2**: **Coverage of causes of deaths and DALYs in the model.** (**A**) The fraction of deaths in the GBD estimates in the period 2015-19 that are due to causes that are mechanistically represented in the model. (**B**) The same as (A) but for DALYs. In each panel, the inlay list the causes that are not mechanistically represented that account for ≥0.5% of all deaths or DALYs in that period.

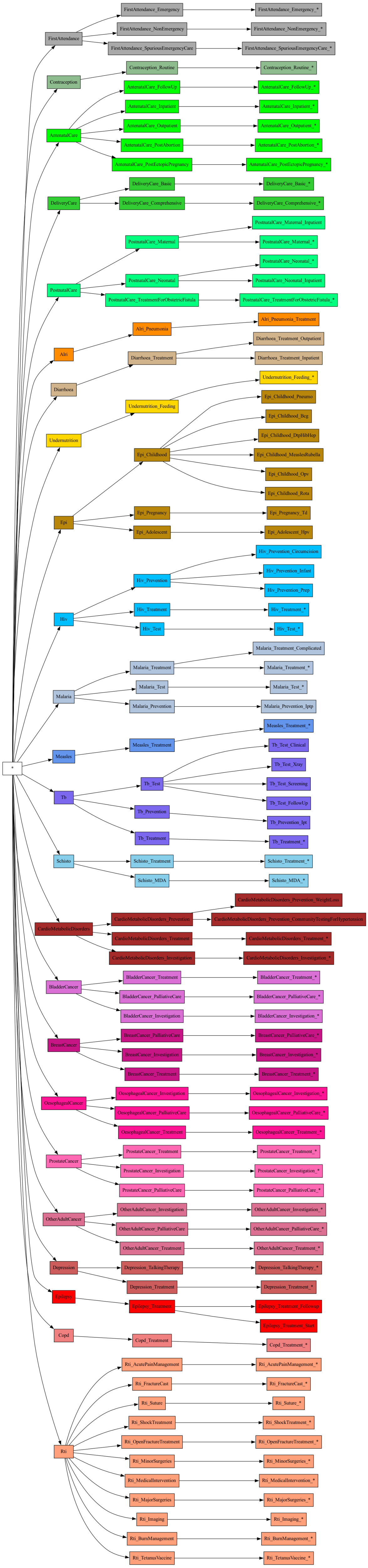

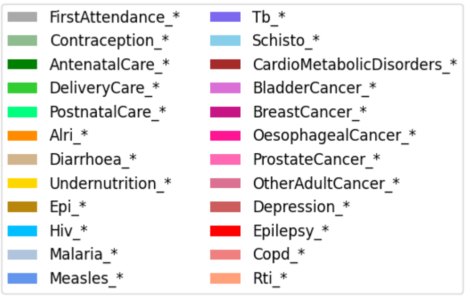


**Figure S3**: The services represented in the model. Each box in the bottom row is an individual service. The other rows show how these are organized into groups that correspond to the care for particular types of condition. The top-most row (and the colour coding) shows the highest level of organization of these services, which corresponds broadly to the main disease/conditions.

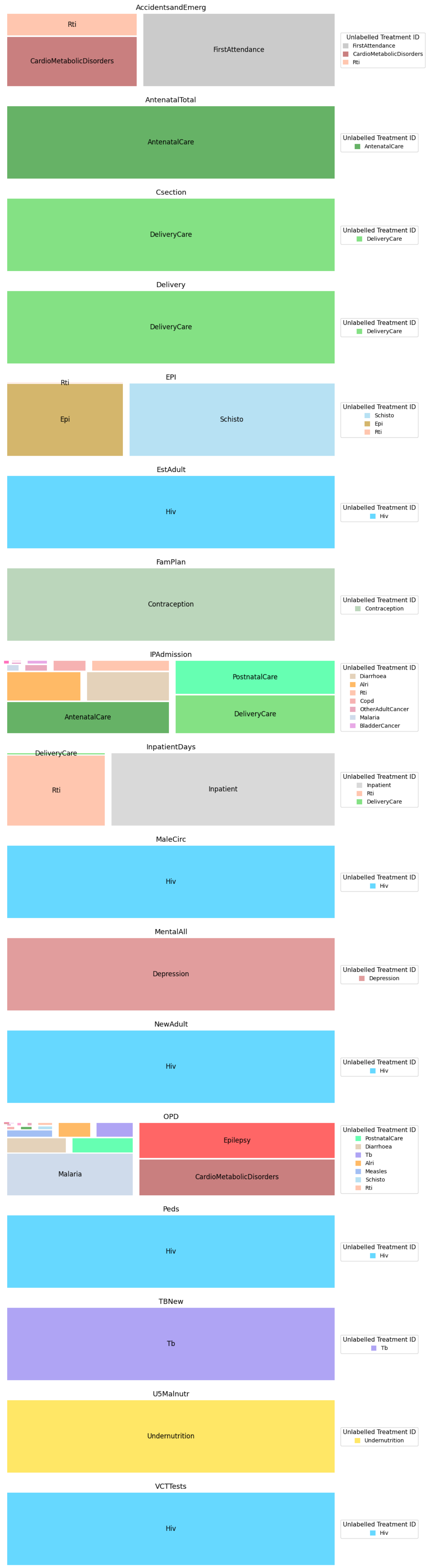


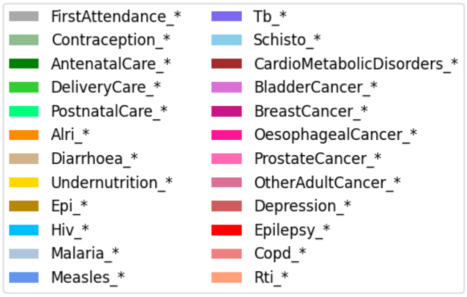


**Figure S4**: The requirement for different types of appointment from each type of healthcare service (“TREATMENT_ID”). Each panel shows, for one type of ‘Appointment’, how demand for this appointment is created by the different healthcare services (indicated in the legends).

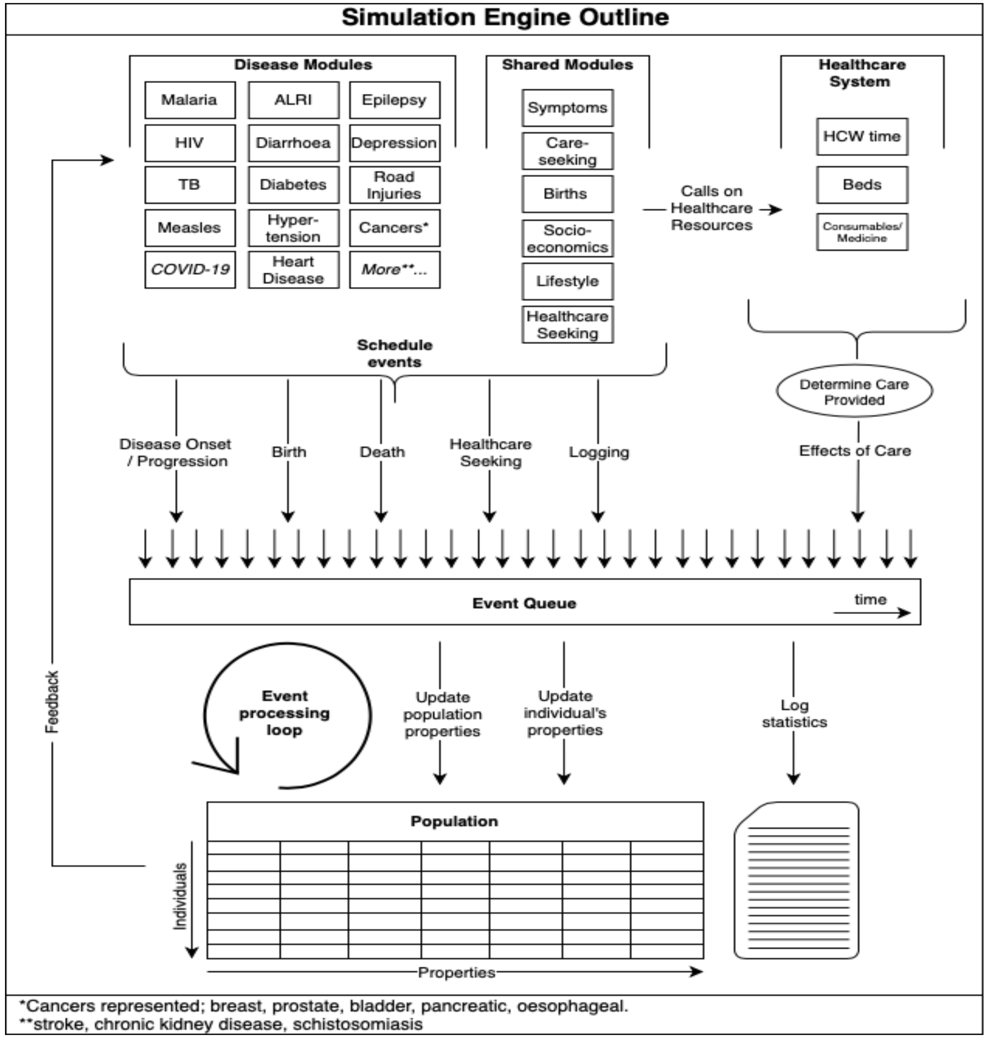


**Figure S5**: **Schematic illustration of the modularized simulation framework.**

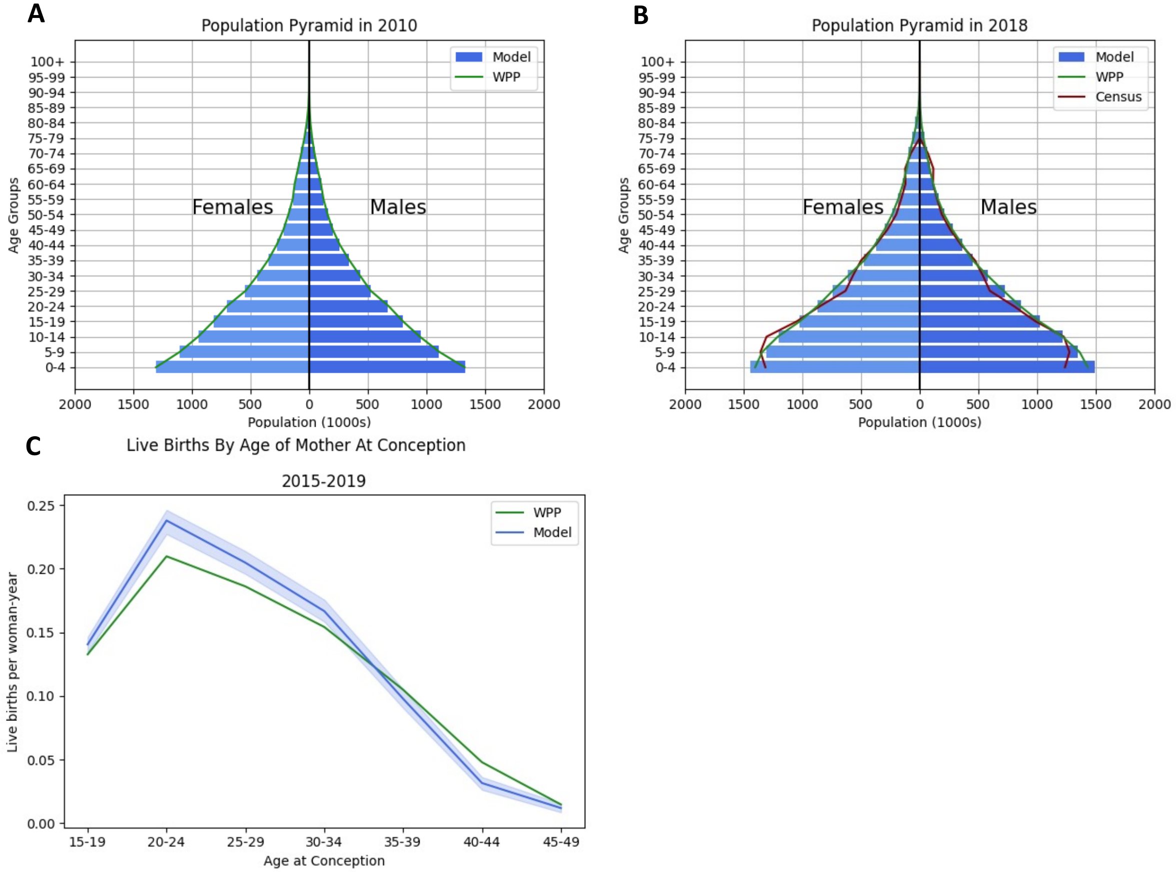


<u>**Figure S6**</u>: **Demographic projections of the model compared to data**. (**A**) The population in 2010 broken down by sex and age-group, in the model compared to the World Population Prospects (WPP). (**B**) The same as (A) but for year 2018 and including the the national census of that year. (**C**) The number of live births per woman per year, by age at conception, in the model and the WPP in the period 2015-19.

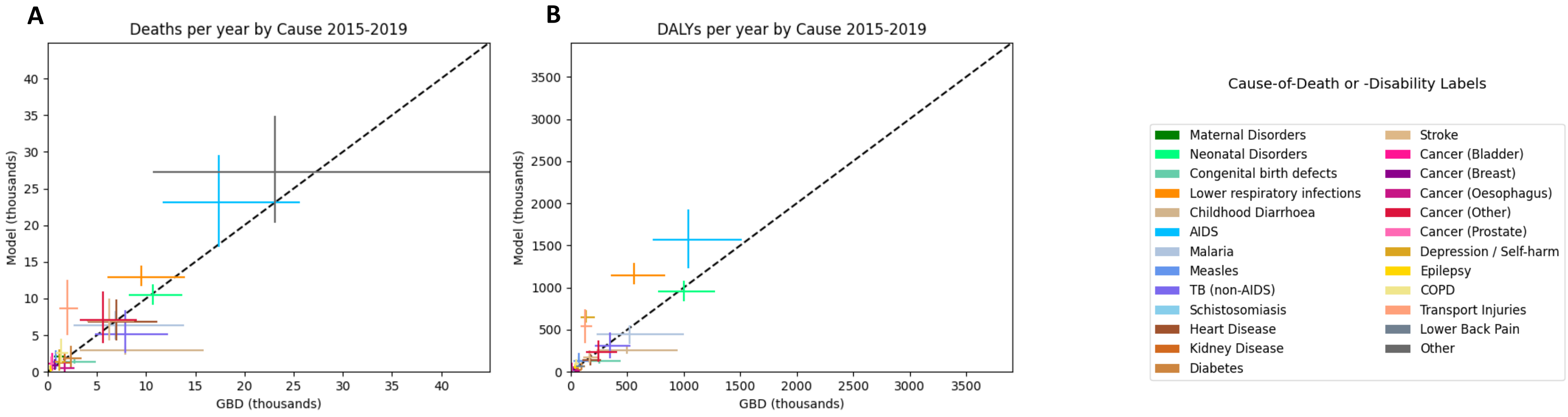


__Figure S7__: **Comparison of the total number of deaths and DALYs by cause between the model and available data**. (**A**) Number of Deaths and (**B**) DALYs incurred for each cause (for persons of all ages) in the period 2015-2019 in the model (vertical axis), compared with the estimates in Global Burden of Disease (GBD) (horizontal axis). The 'Other' cause of death means causes that are not represented mechanistically in this model. The 1:1 line, which indicates equality between the model and GBD estimates, is drawn to aid inspection. The vertical and horizontal error bars display the uncertainty in the model and GBD estimates, respectively.

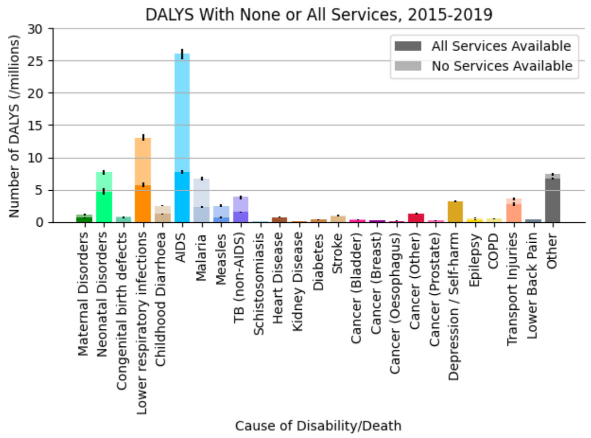


**Figure S8**: **The impact of all healthcare services on the causes of death/disability.** Total DALYs in the period 2015-2019 by cause under the scenario when all services are provided (heavier shading), and when no services are provided (lighter shading).

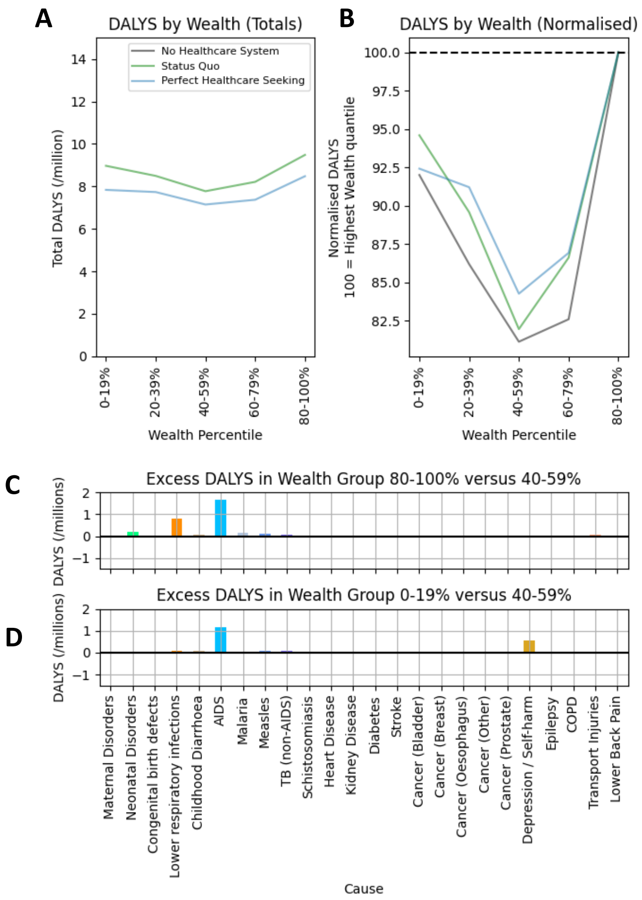


**<u>Figure S9</u>: Distribution of health burden by wealth quintile in the model. (A)** Total DALYS incurred in the period 2015-2019 by wealth quintile under selected scenarios (see Table 1); (**B**) The relative DALY burden under the same scenarios (normalised so that 100 = DALYs of Highest Wealth quintile within each scenario). (**C**) The number of additional DALYS incurred by persons in the highest wealth quintile compared to persons in the lowest wealth quintiles, broken down by cause. (**D**) The number of additional DALYs incurred by persons in the lowest wealth quintile compared to persons in the middle wealth quintiles, broken down by cause.

This shows that the reason for the greater health gains for the highest wealth quintile under the Status Quo scenario (Figure 4(B)) is due to: (i) a greater underlying health burden in the highest wealth quintile of some common diseases that the healthcare system can be treated effectively (i.e., AIDS and Lower Respiratory infections (for which HIV/AIDS is key risk factor for onset); see panel C.) (ii) the greater propensity of persons in higher wealth quintiles to seek care quickly (Ng'ambi *et al.*) (when this difference is removed, the distribution of health burden becomes slightly more evenly distributed: see the comparison in panel B between 'Status Quo' and 'Perfect Healthcare Seeking'). (The reason for the partially "U-shaped" pattern in DALYS averted under Status Quo scenario (Figure 4(B)), is because the lowest wealth quintile also has a higher burden of AIDS than the middle quintile: this is because the distribution of greater HIV is linked to higher wealth (Mangal *et al.*,) but also to lesser educational attainment (Mangal *et al.*,), which is most common in lower wealth quintiles.)

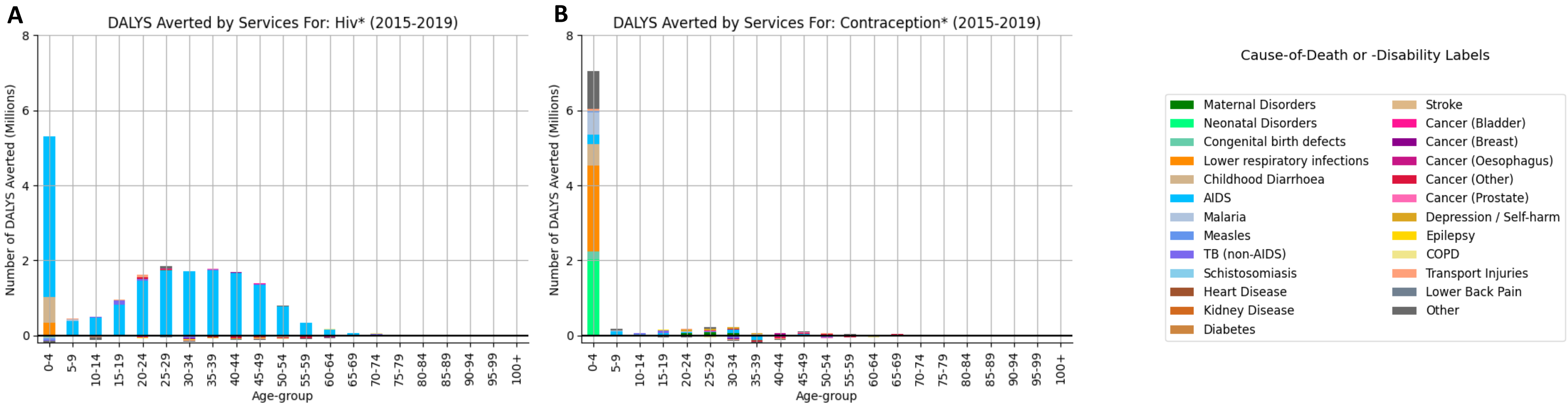


**<u>Figure S10</u>: DALYs averted by specific set of services, broken down by age-group and cause**. (**A**) The DALYs averted by all HIV services; and (**B**) the DALYs averted by all contraception services. The estimate of DALYs averted by a set of services is computed by comparing a simulation in which all services are provided with a simulation in which that specific service is removed.

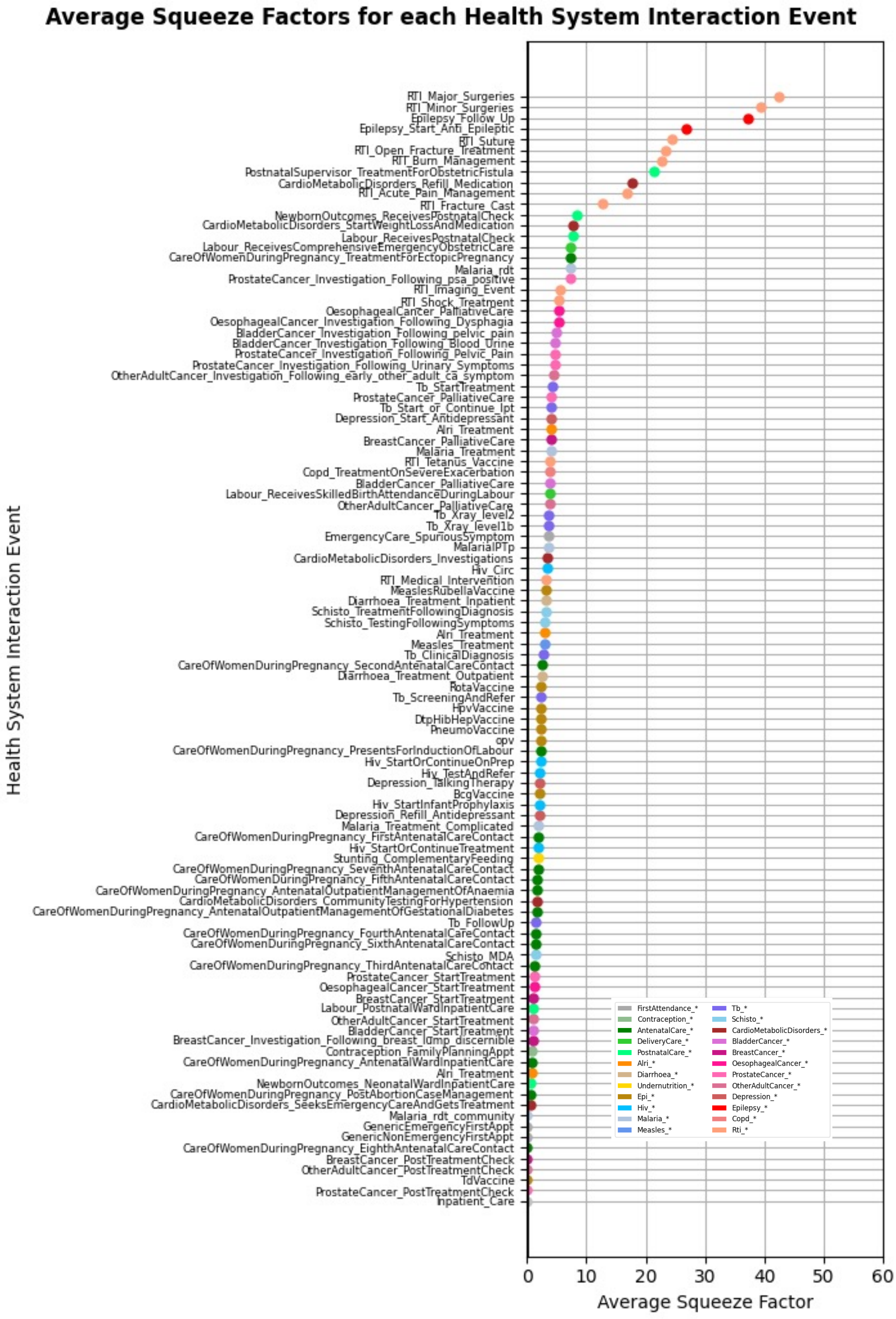


**<u>Figure S11</u>: The Average 'Squeeze-Factor' for each Health System Interaction Event.** The 'squeeze-factor' for each HSI event is computed as: $max_i\left(\frac{c_i}{a_i}-1\right)$, where $c_i$ is total demand for the time of the $i^{th}$ healthcare worker cadre on the day, district and facility-level at which that the HSI occurs, and $a_i$ is the corresponding available time for that healthcare worker cadre. Values greater than 0 indicate that, for at least one healthcare worker cadre that is required in the delivery of that HSI, the total demand for the time of that cadre (at that day and in that facility level) exceeds the time that is available (as per the contracted hours). The average is taken across all occurrences of that event in the period 2015-2019. The names of the HSI event are arranged in order of the average squeeze-factor and the colour of the dot corresponds to the TREATMENT_ID of which they are part.

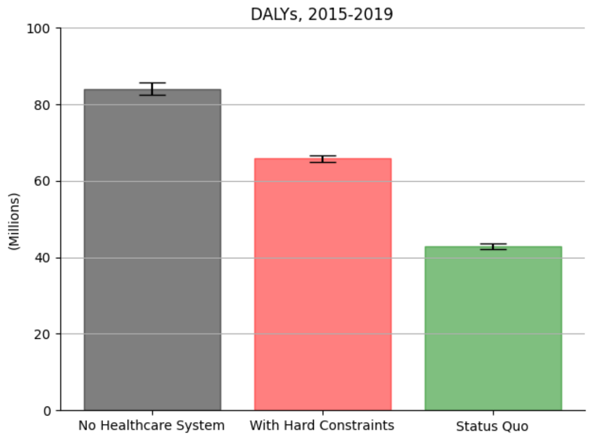


**<u>Figure S12</u>: Health in the population under different assumptions for the healthcare system.** Estimated DALYs incurred in the population in the period 2015-2019 currently (‘Status Quo’: green bar); or, if there we no healthcare provided (grey bar); or, if the ‘Hard Constraints’ assumptions is applied (whereby all appointments take as long as per clinical expectations and there is not healthcare worker over-time; pink bar).

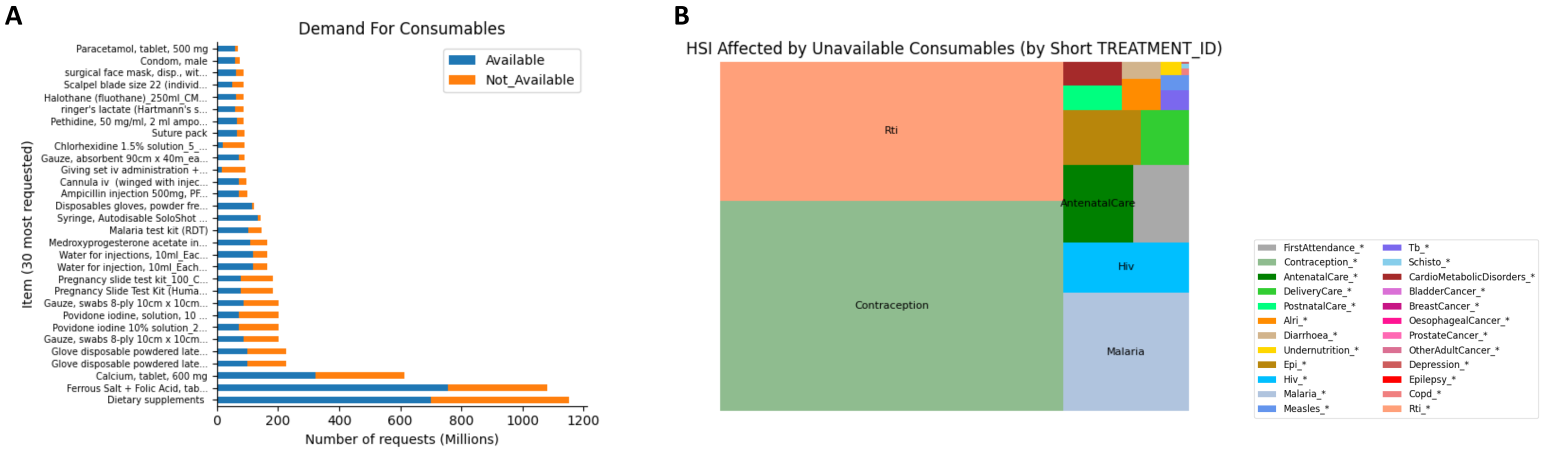


<u>**Figure S13**</u>: **Demand of consumables in the model in the period 2015-19.** (**A**) The number of "requests" for each consumable item in the model, classified by whether the item was available or not at the time and at the facility at which it was requested. Only bars for the twenty most requested consumable items are shown and in descending order from the bottom of the number of requests. (**B**) A breakdown of the healthcare services ("TREATMENT_ID") that are affected by a consumable item not being available, where the area of each segment is proportional to the number of times in which an HSI is run and suffers from a consumable not being available.
