## Supplementary Tables for "A Healthcare Service Delivery and Epidemiological Model for Investigating Resource Allocation for Health: The *Thanzi La Onse* Model"

**<u>Table S1</u>: The Healthcare System Interaction (HSI) events in the model.** Each row is for one HSI and shows the class of HSI it belong to, the service of which that HSI is a part (its "TREATMENT_ID"), the facility level(s) at which it operates and the 'Appointment Type(s)' and 'Bed Type(s)' it requires.

| Module | TREATMENT_ID | HSI Event Class | Facility Level | Appointment Types | Bed-Type |
|---|---|---|---|---|---|
| **Alri** | Alri_Pneumonia_Treatment_Inpatient | Alri_Treatment | 2 | IPAdmission, InpatientDays | general_bed |
| **Alri** | Alri_Pneumonia_Treatment_Inpatient | Alri_Treatment | 2 | - | general_bed |
| **Alri** | Alri_Pneumonia_Treatment_Inpatient_Followup | Alri_Treatment | 1a | IPAdmission, InpatientDays | general_bed |
| **Alri** | Alri_Pneumonia_Treatment_Inpatient_Followup | Alri_Treatment | 2 | IPAdmission, InpatientDays | general_bed |
| **Alri** | Alri_Pneumonia_Treatment_Outpatient | Alri_Treatment | 0 | ConWithDCSA | - |
| **Alri** | Alri_Pneumonia_Treatment_Outpatient | Alri_Treatment | 0 | - | - |
| **Alri** | Alri_Pneumonia_Treatment_Outpatient | Alri_Treatment | 1a | Under5OPD | - |
| **Alri** | Alri_Pneumonia_Treatment_Outpatient | Alri_Treatment | 1a | - | - |
| **Alri** | Alri_Pneumonia_Treatment_Outpatient | Alri_Treatment | 2 | Under5OPD | - |
| **Alri** | Alri_Pneumonia_Treatment_Outpatient | Alri_Treatment | 2 | - | - |
| **BladderCancer** | BladderCancer_Investigation | BladderCancer_Investigation_Following_Blood_Urine | 2 | Over5OPD | - |
| **BladderCancer** | BladderCancer_Investigation | BladderCancer_Investigation_Following_Blood_Urine | 2 | - | - |
| **BladderCancer** | BladderCancer_Investigation | BladderCancer_Investigation_Following_pelvic_pain | 2 | Over5OPD | - |
| **BladderCancer** | BladderCancer_Investigation | BladderCancer_Investigation_Following_pelvic_pain | 2 | - | - |
| **BladderCancer** | BladderCancer_PalliativeCare | BladderCancer_PalliativeCare | 2 | IPAdmission, InpatientDays | general_bed |
| **BladderCancer** | BladderCancer_Treatment | BladderCancer_PostTreatmentCheck | 3 | Over5OPD | - |

| | | | | | |
|---|---|---|---|---|---|
| **BladderCancer** | BladderCancer_Treatment | BladderCancer_StartTreatment | 3 | IPAdmission, InpatientDays, MajorSurg | general_bed |
| **BreastCancer** | BreastCancer_Investigation | BreastCancer_Investigation_Following_breast_lump_discernible | 3 | - | - |
| **BreastCancer** | BreastCancer_Investigation | BreastCancer_Investigation_Following_breast_lump_discernible | 3 | Mammography, Over5OPD | - |
| **BreastCancer** | BreastCancer_PalliativeCare | BreastCancer_PalliativeCare | 2 | IPAdmission, InpatientDays | general_bed |
| **BreastCancer** | BreastCancer_Treatment | BreastCancer_PostTreatmentCheck | 3 | Over5OPD | - |
| **BreastCancer** | BreastCancer_Treatment | BreastCancer_StartTreatment | 3 | IPAdmission, InpatientDays, MajorSurg | general_bed |
| **CardioMetabolicDisorders** | CardioMetabolicDisorders_Investigation | CardioMetabolicDisorders_Investigations | 2 | Over5OPD | - |
| **CardioMetabolicDisorders** | CardioMetabolicDisorders_Prevention_CommunityTestingForHypertension | CardioMetabolicDisorders_CommunityTestingForHypertension | 1a | Over5OPD | - |
| **CardioMetabolicDisorders** | CardioMetabolicDisorders_Prevention_WeightLoss | CardioMetabolicDisorders_StartWeightLossAndMedication | 2 | Over5OPD | - |
| **CardioMetabolicDisorders** | CardioMetabolicDisorders_Prevention_WeightLoss | CardioMetabolicDisorders_StartWeightLossAndMedication | 2 | - | - |
| **CardioMetabolicDisorders** | CardioMetabolicDisorders_Treatment | CardioMetabolicDisorders_Refill_Medication | 2 | Over5OPD | - |
| **CardioMetabolicDisorders** | CardioMetabolicDisorders_Treatment | CardioMetabolicDisorders_Refill_Medication | 2 | - | - |
| **CardioMetabolicDisorders** | CardioMetabolicDisorders_Treatment | CardioMetabolicDisorders_SeeksEmergencyCareAndGetsTreatment | 2 | AccidentsandEmerg | - |
| **CareOfWomenDuringPregnancy** | AntenatalCare_FollowUp | CareOfWomenDuringPregnancy_AntenatalOutpatientManagementOfAnaemia | 1a | Over5OPD | - |

| | | | | | |
|---|---|---|---|---|---|
| **CareOfWomenDuringPregnancy** | AntenatalCare_FollowUp | CareOfWomenDuringPregnancy_AntenatalOutpatientManagementOfGestationalDiabetes | 1a | Over5OPD | - |
| **CareOfWomenDuringPregnancy** | AntenatalCare_Inpatient | CareOfWomenDuringPregnancy_AntenatalWardInpatientCare | 2 | IPAdmission, InpatientDays | maternity_bed |
| **CareOfWomenDuringPregnancy** | AntenatalCare_Inpatient | CareOfWomenDuringPregnancy_PresentsForInductionOfLabour | 1a | IPAdmission, InpatientDays | maternity_bed |
| **CareOfWomenDuringPregnancy** | AntenatalCare_Outpatient | CareOfWomenDuringPregnancy_FifthAntenatalCareContact | 1a | ANCSubsequent | - |
| **CareOfWomenDuringPregnancy** | AntenatalCare_Outpatient | CareOfWomenDuringPregnancy_FifthAntenatalCareContact | 1a | - | - |
| **CareOfWomenDuringPregnancy** | AntenatalCare_Outpatient | CareOfWomenDuringPregnancy_FirstAntenatalCareContact | 1a | AntenatalFirst | - |
| **CareOfWomenDuringPregnancy** | AntenatalCare_Outpatient | CareOfWomenDuringPregnancy_FirstAntenatalCareContact | 1a | - | - |
| **CareOfWomenDuringPregnancy** | AntenatalCare_Outpatient | CareOfWomenDuringPregnancy_FourthAntenatalCareContact | 1a | ANCSubsequent | - |
| **CareOfWomenDuringPregnancy** | AntenatalCare_Outpatient | CareOfWomenDuringPregnancy_FourthAntenatalCareContact | 1a | - | - |
| **CareOfWomenDuringPregnancy** | AntenatalCare_Outpatient | CareOfWomenDuringPregnancy_SecondAntenatalCareContact | 1a | ANCSubsequent | - |
| **CareOfWomenDuringPregnancy** | AntenatalCare_Outpatient | CareOfWomenDuringPregnancy_SecondAntenatalCareContact | 1a | - | - |
| **CareOfWomenDuringPregnancy** | AntenatalCare_Outpatient | CareOfWomenDuringPregnancy_SeventhAntenatalCareContact | 1a | - | - |
| **CareOfWomenDuringPregnancy** | AntenatalCare_Outpatient | CareOfWomenDuringPregnancy_SeventhAntenatalCareContact | 1a | ANCSubsequent | - |
| **CareOfWomenDuringPregnancy** | AntenatalCare_Outpatient | CareOfWomenDuringPregnancy_SixthAntenatalCareContact | 1a | ANCSubsequent | - |

| | | | | | |
|---|---|---|---|---|---|
| **CareOfWomenDuringPregnancy** | AntenatalCare_Outpatient | CareOfWomenDuringPregnancy_SixthAntenatalCareContact | 1a | - | - |
| **CareOfWomenDuringPregnancy** | AntenatalCare_Outpatient | CareOfWomenDuringPregnancy_ThirdAntenatalCareContact | 1a | ANCSubsequent | - |
| **CareOfWomenDuringPregnancy** | AntenatalCare_Outpatient | CareOfWomenDuringPregnancy_ThirdAntenatalCareContact | 1a | - | - |
| **CareOfWomenDuringPregnancy** | AntenatalCare_PostAbortion | CareOfWomenDuringPregnancy_PostAbortionCaseManagement | 2 | IPAdmission, InpatientDays | maternity_bed |
| **CareOfWomenDuringPregnancy** | AntenatalCare_PostEctopicPregnancy | CareOfWomenDuringPregnancy_TreatmentForEctopicPregnancy | 2 | IPAdmission, InpatientDays, MajorSurg | maternity_bed |
| **Contraception** | Contraception_Routine | Contraception_FamilyPlanningAppt | 1a | FamPlan | - |
| **Contraception** | Contraception_Routine | Contraception_FamilyPlanningAppt | 1a | - | - |
| **Contraception** | Contraception_Routine | Contraception_FamilyPlanningAppt | 1a | PharmDispensing | - |
| **Contraception** | Contraception_Routine | Contraception_FamilyPlanningAppt | 2 | FamPlan | - |
| **Contraception** | Contraception_Routine | Contraception_FamilyPlanningAppt | 2 | - | - |
| **Contraception** | Contraception_Routine | Contraception_FamilyPlanningAppt | 2 | MinorSurg | - |
| **Depression** | Depression_TalkingTherapy | Depression_TalkingTherapy | 2 | MentOPD | - |
| **Depression** | Depression_Treatment | Depression_Refill_Antidepressant | 1a | - | - |
| **Depression** | Depression_Treatment | Depression_Refill_Antidepressant | 1a | Over5OPD | - |
| **Depression** | Depression_Treatment | Depression_Start_Antidepressant | 2 | MentOPD | - |
| **Depression** | Depression_Treatment | Depression_Start_Antidepressant | 2 | - | - |
| **Diarrhoea** | Diarrhoea_Treatment_Inpatient | Diarrhoea_Treatment_Inpatient | 1a | IPAdmission, InpatientDays | general_bed |
| **Diarrhoea** | Diarrhoea_Treatment_Outpatient | Diarrhoea_Treatment_Outpatient | 1a | Under5OPD | - |

| | | | | | |
|---|---|---|---|---|---|
| **Epi** | Epi_Adolescent_Hpv | HpvVaccine | 1a | EPI | - |
| **Epi** | Epi_Adolescent_Hpv | HpvVaccine | 2 | EPI | - |
| **Epi** | Epi_Childhood_Bcg | BcgVaccine | 1a | EPI | - |
| **Epi** | Epi_Childhood_Bcg | BcgVaccine | 2 | EPI | - |
| **Epi** | Epi_Childhood_DtpHibHep | DtpHibHepVaccine | 1a | EPI | - |
| **Epi** | Epi_Childhood_DtpHibHep | DtpHibHepVaccine | 2 | EPI | - |
| **Epi** | Epi_Childhood_MeaslesRubella | MeaslesRubellaVaccine | 1a | EPI | - |
| **Epi** | Epi_Childhood_MeaslesRubella | MeaslesRubellaVaccine | 2 | EPI | - |
| **Epi** | Epi_Childhood_Opv | opv | 1a | EPI | - |
| **Epi** | Epi_Childhood_Opv | opv | 2 | EPI | - |
| **Epi** | Epi_Childhood_Pneumo | PneumoVaccine | 1a | EPI | - |
| **Epi** | Epi_Childhood_Pneumo | PneumoVaccine | 2 | EPI | - |
| **Epi** | Epi_Childhood_Rota | RotaVaccine | 1a | EPI | - |
| **Epi** | Epi_Childhood_Rota | RotaVaccine | 2 | EPI | - |
| **Epi** | Epi_Pregnancy_Td | TdVaccine | 1a | - | - |
| **Epilepsy** | Epilepsy_Treatment_Followup | Epilepsy_Follow_Up | 2 | Over5OPD | - |
| **Epilepsy** | Epilepsy_Treatment_Followup | Epilepsy_Follow_Up | 2 | - | - |
| **Epilepsy** | Epilepsy_Treatment_Followup | Epilepsy_Follow_Up | 2 | PharmDispensing | - |
| **Epilepsy** | Epilepsy_Treatment_Start | Epilepsy_Start_Anti_Epileptic | 2 | Over5OPD | - |
| **HealthSeekingBehaviour** | FirstAttendance_Emergency | GenericEmergencyFirstAppt | 2 | - | - |
| **HealthSeekingBehaviour** | FirstAttendance_NonEmergency | GenericNonEmergencyFirstAppt | 0 | - | - |
| **HealthSeekingBehaviour** | FirstAttendance_NonEmergency | GenericNonEmergencyFirstAppt | 1a | - | - |
| **HealthSeekingBehaviour** | FirstAttendance_NonEmergency | GenericNonEmergencyFirstAppt | 2 | - | - |
| **HealthSeekingBehaviour** | FirstAttendance_SpuriousEmergencyCare | EmergencyCare_SpuriousSymptom | 1a | AccidentsandEmerg | - |

| | | | | | |
|---|---|---|---|---|---|
| **HealthSystem** | Inpatient_Care | Inpatient_Care | 3 | InpatientDays | - |
| **Hiv** | Hiv_Prevention_Circumcision | Hiv_Circ | 1a | MaleCirc | - |
| **Hiv** | Hiv_Prevention_Infant | Hiv_StartInfantProphylaxis | 1a | - | - |
| **Hiv** | Hiv_Prevention_Infant | Hiv_StartInfantProphylaxis | 1a | Peds, VCTNegative | - |
| **Hiv** | Hiv_Prevention_Prep | Hiv_StartOrContinueOnPrep | 1a | PharmDispensing, VCTNegative | - |
| **Hiv** | Hiv_Test | Hiv_TestAndRefer | 1a | VCTPositive | - |
| **Hiv** | Hiv_Test | Hiv_TestAndRefer | 1a | VCTNegative | - |
| **Hiv** | Hiv_Test | Hiv_TestAndRefer | 1a | - | - |
| **Hiv** | Hiv_Treatment | Hiv_StartOrContinueTreatment | 1a | NewAdult | - |
| **Hiv** | Hiv_Treatment | Hiv_StartOrContinueTreatment | 1a | Peds | - |
| **Hiv** | Hiv_Treatment | Hiv_StartOrContinueTreatment | 1a | EstNonCom | - |
| **Labour** | DeliveryCare_Basic | Labour_ReceivesSkilledBirthAttendanceDuringLabour | 1a | IPAdmission, InpatientDays, NormalDelivery | maternity_bed |
| **Labour** | DeliveryCare_Basic | Labour_ReceivesSkilledBirthAttendanceDuringLabour | 1a | CompDelivery | maternity_bed |
| **Labour** | DeliveryCare_Basic | Labour_ReceivesSkilledBirthAttendanceDuringLabour | 2 | IPAdmission, InpatientDays, NormalDelivery | maternity_bed |
| **Labour** | DeliveryCare_Basic | Labour_ReceivesSkilledBirthAttendanceDuringLabour | 2 | CompDelivery | maternity_bed |
| **Labour** | DeliveryCare_Comprehensive | Labour_ReceivesComprehensiveEmergencyObstetricCare | 2 | Csection | - |
| **Labour** | DeliveryCare_Comprehensive | Labour_ReceivesComprehensiveEmergencyObstetricCare | 2 | InpatientDays | - |
| **Labour** | DeliveryCare_Comprehensive | Labour_ReceivesComprehensiveEmergencyObstetricCare | 2 | MajorSurg | - |
| **Labour** | FirstAttendance_Emergency | GenericEmergencyFirstAppt | 2 | - | - |

| | | | | | |
|---|---|---|---|---|---|
| **Labour** | PostnatalCare_Maternal | Labour_ReceivesPostnatalCheck | 1a | Over5OPD | - |
| **Labour** | PostnatalCare_Maternal | Labour_ReceivesPostnatalCheck | 2 | Over5OPD | - |
| **Labour** | PostnatalCare_Maternal_Inpatient | Labour_PostnatalWardInpatientCare | 2 | IPAdmission, InpatientDays | maternity_bed |
| **Malaria** | Malaria_Prevention_Iptp | MalarialPTp | 1a | Over5OPD | - |
| **Malaria** | Malaria_Test | Malaria_rdt | 1a | Over5OPD | - |
| **Malaria** | Malaria_Test | Malaria_rdt | 1a | Under5OPD | - |
| **Malaria** | Malaria_Test | Malaria_rdt | 1a | - | - |
| **Malaria** | Malaria_Test | Malaria_rdt | 2 | Over5OPD | - |
| **Malaria** | Malaria_Test | Malaria_rdt | 2 | Under5OPD | - |
| **Malaria** | Malaria_Test | Malaria_rdt | 2 | - | - |
| **Malaria** | Malaria_Test | Malaria_rdt_community | 0 | ConWithDCSA | - |
| **Malaria** | Malaria_Test | Malaria_rdt_community | 0 | - | - |
| **Malaria** | Malaria_Treatment | Malaria_Treatment | 1a | Under5OPD | - |
| **Malaria** | Malaria_Treatment | Malaria_Treatment | 1a | Over5OPD | - |
| **Malaria** | Malaria_Treatment_Complicated | Malaria_Treatment_Complicated | 2 | IPAdmission, InpatientDays, Under5OPD | general_bed |
| **Malaria** | Malaria_Treatment_Complicated | Malaria_Treatment_Complicated | 2 | IPAdmission, InpatientDays, Over5OPD | general_bed |
| **Measles** | Measles_Treatment | Measles_Treatment | 1a | Over5OPD | - |
| **Measles** | Measles_Treatment | Measles_Treatment | 1a | Under5OPD | - |
| **NewbornOutcomes** | PostnatalCare_Neonatal | NewbornOutcomes_ReceivesPostnatalCheck | 1a | Under5OPD | - |
| **NewbornOutcomes** | PostnatalCare_Neonatal | NewbornOutcomes_ReceivesPostnatalCheck | 2 | Under5OPD | - |
| **NewbornOutcomes** | PostnatalCare_Neonatal_Inpatient | NewbornOutcomes_NeonatalWardInpatientCare | 2 | IPAdmission, InpatientDays | general_bed |

| | | | | | |
|---|---|---|---|---|---|
| **OesophagealCancer** | OesophagealCancer_Investigation | OesophagealCancer_Investigation_Following_Dysphagia | 2 | Over5OPD | - |
| **OesophagealCancer** | OesophagealCancer_Investigation | OesophagealCancer_Investigation_Following_Dysphagia | 2 | - | - |
| **OesophagealCancer** | OesophagealCancer_PalliativeCare | OesophagealCancer_PalliativeCare | 2 | IPAdmission, InpatientDays | general_bed |
| **OesophagealCancer** | OesophagealCancer_Treatment | OesophagealCancer_PostTreatmentCheck | 3 | Over5OPD | - |
| **OesophagealCancer** | OesophagealCancer_Treatment | OesophagealCancer_StartTreatment | 3 | IPAdmission, InpatientDays, MajorSurg | general_bed |
| **OtherAdultCancer** | OtherAdultCancer_Investigation | OtherAdultCancer_Investigation_Following_early_other_adult_ca_symptom | 2 | Over5OPD | - |
| **OtherAdultCancer** | OtherAdultCancer_Investigation | OtherAdultCancer_Investigation_Following_early_other_adult_ca_symptom | 2 | - | - |
| **OtherAdultCancer** | OtherAdultCancer_PalliativeCare | OtherAdultCancer_PalliativeCare | 2 | IPAdmission, InpatientDays | general_bed |
| **OtherAdultCancer** | OtherAdultCancer_Treatment | OtherAdultCancer_PostTreatmentCheck | 3 | Over5OPD | - |
| **OtherAdultCancer** | OtherAdultCancer_Treatment | OtherAdultCancer_StartTreatment | 3 | IPAdmission, InpatientDays, MajorSurg | general_bed |
| **PostnatalSupervisor** | PostnatalCare_TreatmentForObstetricFistula | PostnatalSupervisor_TreatmentForObstetricFistula | 2 | IPAdmission, InpatientDays, MajorSurg | general_bed |
| **PregnancySupervisor** | FirstAttendance_Emergency | GenericEmergencyFirstAppt | 2 | - | - |
| **ProstateCancer** | ProstateCancer_Investigation | ProstateCancer_Investigation_Following_Pelvic_Pain | 2 | - | - |
| **ProstateCancer** | ProstateCancer_Investigation | ProstateCancer_Investigation_Following_Pelvic_Pain | 2 | Over5OPD | - |
| **ProstateCancer** | ProstateCancer_Investigation | ProstateCancer_Investigation_Following_Urinary_Symptoms | 2 | - | - |

| | | | | | |
|---|---|---|---|---|---|
| **ProstateCancer** | ProstateCancer_Investigation | ProstateCancer_Investigation_Following_Urinary_Symptoms | 2 | Over5OPD | - |
| **ProstateCancer** | ProstateCancer_Investigation | ProstateCancer_Investigation_Following_psa_positive | 2 | Over5OPD | - |
| **ProstateCancer** | ProstateCancer_Investigation | ProstateCancer_Investigation_Following_psa_positive | 2 | - | - |
| **ProstateCancer** | ProstateCancer_PalliativeCare | ProstateCancer_PalliativeCare | 2 | IPAdmission, InpatientDays | general_bed |
| **ProstateCancer** | ProstateCancer_Treatment | ProstateCancer_PostTreatmentCheck | 3 | Over5OPD | - |
| **ProstateCancer** | ProstateCancer_Treatment | ProstateCancer_StartTreatment | 3 | IPAdmission, InpatientDays, MajorSurg | general_bed |
| **RTI** | Rti_AcutePainManagement | RTI_Acute_Pain_Management | 2 | Over5OPD | - |
| **RTI** | Rti_AcutePainManagement | RTI_Acute_Pain_Management | 2 | - | - |
| **RTI** | Rti_AcutePainManagement | RTI_Acute_Pain_Management | 2 | Under5OPD | - |
| **RTI** | Rti_BurnManagement | RTI_Burn_Management | 2 | IPAdmission, InpatientDays, MinorSurg | general_bed |
| **RTI** | Rti_FractureCast | RTI_Fracture_Cast | 2 | AccidentsandEmerg | - |
| **RTI** | Rti_FractureCast | RTI_Fracture_Cast | 2 | - | - |
| **RTI** | Rti_Imaging | RTI_Imaging_Event | 2 | DiagRadio | - |
| **RTI** | Rti_Imaging | RTI_Imaging_Event | 3 | DiagRadio, Tomography | - |
| **RTI** | Rti_MajorSurgeries | RTI_Major_Surgeries | 2 | - | - |
| **RTI** | Rti_MajorSurgeries | RTI_Major_Surgeries | 2 | MajorSurg | - |
| **RTI** | Rti_MedicalIntervention | RTI_Medical_Intervention | 2 | AccidentsandEmerg, IPAdmission, InpatientDays | general_bed |
| **RTI** | Rti_MedicalIntervention | RTI_Medical_Intervention | 2 | AccidentsandEmerg, IPAdmission, InpatientDays | - |
| **RTI** | Rti_MinorSurgeries | RTI_Minor_Surgeries | 2 | - | - |

| | | | | | |
|---|---|---|---|---|---|
| **RTI** | Rti_MinorSurgeries | RTI_Minor_Surgeries | 2 | MinorSurg | - |
| **RTI** | Rti_OpenFractureTreatment | RTI_Open_Fracture_Treatment | 2 | MinorSurg | - |
| **RTI** | Rti_ShockTreatment | RTI_Shock_Treatment | 2 | - | - |
| **RTI** | Rti_ShockTreatment | RTI_Shock_Treatment | 2 | AccidentsandEmerg | - |
| **RTI** | Rti_Suture | RTI_Suture | 2 | - | - |
| **RTI** | Rti_Suture | RTI_Suture | 2 | Over5OPD | - |
| **RTI** | Rti_Suture | RTI_Suture | 2 | Under5OPD | - |
| **RTI** | Rti_TetanusVaccine | RTI_Tetanus_Vaccine | 2 | EPI | - |
| **RTI** | Rti_TetanusVaccine | RTI_Tetanus_Vaccine | 2 | - | - |
| **Schisto** | Schisto_MDA | Schisto_MDA | 1a | EPI | - |
| **Schisto** | Schisto_Treatment | Schisto_TestingFollowingSymptoms | 1a | Over5OPD | - |
| **Schisto** | Schisto_Treatment | Schisto_TestingFollowingSymptoms | 1a | Under5OPD | - |
| **Schisto** | Schisto_Treatment | Schisto_TreatmentFollowingDiagnosis | 1a | Under5OPD | - |
| **Schisto** | Schisto_Treatment | Schisto_TreatmentFollowingDiagnosis | 1a | Over5OPD | - |
| **Stunting** | Undernutrition_Feeding | Stunting_ComplementaryFeeding | 1a | U5Malnutr | - |
| **Tb** | Tb_Prevention_Ipt | Tb_Start_or_Continue_Ipt | 1a | Over5OPD | - |
| **Tb** | Tb_Test_FollowUp | Tb_FollowUp | 1a | LabTBMicro, TBFollowUp | - |
| **Tb** | Tb_Test_FollowUp | Tb_FollowUp | 1a | LabMolec, LabTBMicro, TBFollowUp | - |
| **Tb** | Tb_Test_FollowUp | Tb_FollowUp | 1a | TBFollowUp | - |
| **Tb** | Tb_Test_Screening | Tb_ScreeningAndRefer | 1a | - | - |
| **Tb** | Tb_Test_Screening | Tb_ScreeningAndRefer | 1a | LabTBMicro, Over5OPD | - |

| | | | | | |
|---|---|---|---|---|---|
| **Tb** | Tb_Test_Screening | Tb_ScreeningAndRefer | 1a | Under5OPD | - |
| **Tb** | Tb_Test_Xray | Tb_Xray_level1b | 2 | DiagRadio | - |
| **Tb** | Tb_Test_Xray | Tb_Xray_level1b | 2 | DiagRadio, Under5OPD | - |
| **Tb** | Tb_Test_Xray | Tb_Xray_level2 | 2 | DiagRadio, Under5OPD | - |
| **Tb** | Tb_Test_Xray | Tb_Xray_level2 | 2 | DiagRadio | - |
| **Tb** | Tb_Test_Xray | Tb_Xray_level2 | 2 | - | - |
| **Tb** | Tb_Treatment | Tb_StartTreatment | 1a | TBNew | - |
| **Tb** | Tb_Treatment | Tb_StartTreatment | 1a | PharmDispensing | - |
| **Tb** | Tb_Treatment | Tb_StartTreatment | 1a | - | - |
